# Neonatal mortality risk of large-for-gestational age and macrosomic live births in low- and middle-income subnational birth cohorts: An individual participant meta-analysis (2000-2017)

**DOI:** 10.64898/2026.06.03.26354851

**Authors:** Fati Kirakoya-Samadoulougou, Blaise Barche, Joyeuse Ukwishaka, Seema Subedi, Daniel J. Erchick, Lorena Suarez Idueta, Davidson H. Hamer, Katherine E.A. Semrau, Fern Mweene Hamomba, Bowen Banda, Albert Manasyan, Jake M. Pry, Kenneth Maleta, Ulla Ashorn, Christentze Schmiegelow, Line Hjort, Daniel T.R. Minja, John P.A. Lusingu, Mariângela Freitas da Silveira, Romina Buffarini, Abdullah H. Baqui, Rasheda Khanam, Salahuddin Ahmed, Zhonghai Zhu, Lingxia Zeng, Yue Cheng, Carl Lachat, Dominique Roberfroid, Lieven Huybregts, Laéticia Céline Toe, James M. Tielsch, Subarna K Khatry, Luke C. Mullany, Eric O. Ohuma, Hannah Blencowe, Joanne Katz, Anne C.C. Lee, Robert Black, Elizabeth A. Hazel

## Abstract

**Background:** Large-for-gestational-age (LGA) and macrosomic newborns are at increased risk of adverse perinatal outcomes, including death, yet the burden of neonatal mortality associated with these conditions in low- and middle-income countries (LMICs), where ongoing nutritional and epidemiological transitions suggest their prevalence will rise, remains poorly quantified. In this study, we quantify the neonatal mortality risk associated with LGA and macrosomia from 16 subnational birth cohorts in low- and middle-income countries between 2000 and 2017.

**Methods and findings:** This is an individual-participant meta-analysis to estimate neonatal mortality rates (NMRs) and relative risks among LGA infants (>90th and >97th percentile birth weight-for-gestational-age using INTERGROWTH-21st) versus appropriate-for-gestational-age (AGA, 10th-90th percentile) infants. Macrosomic (≥4000 g and ≥4500 g) neonates were compared with those weighing 2500 g-3999g. Missing birth weights were imputed using recalibration and multiple imputation methods. We used random effects meta-analysis to pool relative risks.

Median prevalences of LGA >90th and >97th percentile were 5.3% (interquartile range 3.6-8.2) and 2.6% (IQR 1.3-4.5), respectively; macrosomia (≥4000 g and ≥4500 g) prevalences were 1.0% (IQR 0.3-3.1) and 0.06% (IQR 0.0, 0.30), respectively. Mortality was highest among preterm plus LGA infants (61.3 per 1000). LGA infants in the >90th percentile had over twofold increased mortality compared with appropriate-for-gestational-age infants (RR: 2.46; 95% CI: 1.86-3.25), while >97th percentile infants had a higher risk (RR: 3.77; 95% CI: 2.50-5.69). Term LGA >97th percentile infants also showed elevated mortality (RR: 3.14; 95% CI: 1.58-6.22). For LGA >97th percentile, the risk was higher in the early neonatal period (RR: 2.71; 95% CI: 1.92-3.82) than late (RR: 1.69; 95% CI: 1.22-2.34). There was no overall association between macrosomia (≥4000 g) and neonatal mortality. Population attributable fractions were 7.2% for LGA >90th percentile and 0.4% for macrosomia (≥4000 g).

**Conclusions:** Neonatal mortality risks were elevated among LGA infants in low- and middle-income countries, particularly at extreme values (>97th percentile) and during the early neonatal period. Macrosomia showed weaker, less robust associations. Although LGA prevalence is currently low (∼5%) and contributes less to neonatal mortality than small newborns, ongoing nutritional and epidemiological transitions suggest increasing prevalence. This highlights the need for strengthened surveillance, monitoring, and improved delivery planning to ensure that no population is left behind.

## INTRODUCTION

In 2022, an estimated 2.3 million newborns died within the first 28 days of life, more than 80% of them in low- and middle-income countries (LMICs) (1). Sub-Saharan Africa had the highest neonatal mortality rate (NMR) at 27 deaths per 1000 live births, followed by Central and South Asia at 21 deaths per 1000 live births (1). While neonatal mortality interventions have focused on small vulnerable newborns (2), those born preterm, small-for-gestational-age, or with low birth weight, recent advances in neonatal classification, which integrate gestational age and size-for-gestational-age, underscore the need to consider risks across the full spectrum of newborn size (3). Large-for-gestational-age (LGA; >90th percentile for gestational age and sex according to a standard population) and macrosomic newborns (≥4000 g) also face risks of adverse perinatal outcomes, including neonatal death (3–8). Addressing both extremes of birth weight and gestational age is therefore essential for reducing neonatal mortality, particularly in LMICs, where the burden of adverse outcomes is greatest and clinical interventions are more limited (3,5)

In LMICs, the median prevalence of LGA and macrosomia is relatively low, at 5.1% and 1.3%, respectively, compared with approximately 20% and 10% in high-income countries (HICs) (3,9). LGA prevalence in healthy populations is expected to be ∼10% owing to natural variation; levels above this threshold may indicate poor population-level health commonly associated with maternal hyperglycemia, obesity, or excessive gestational weight gain, which increases the risk of fetal overgrowth (10,11). LGA and macrosomic births are associated with increased risks of obstructed labor, birth asphyxia, birth injuries, and cesarean delivery—complications that carry heightened consequences in LMIC settings where access to emergency obstetric care and neonatal resuscitation is often limited. Moreover, the rising prevalence of maternal obesity, gestational diabetes, and overnutrition in LMICs due to ongoing nutritional and epidemiological transitions suggests that LGA and macrosomia will become an increasingly significant public health concern.

Existing analyses, largely from high- and upper-middle-income countries, report inconsistent findings regarding the contribution of LGA to neonatal mortality. For instance, some studies report no significant association between LGA and neonatal mortality (5,12), whereas others have identified greater morbidity among term LGA infants than among appropriate-for-gestational-age (AGA) infants, despite similar rates of stillbirth and neonatal mortality (13). Conversely, Suárez-Idueta reported that neonatal mortality rates are lower among LGA newborns (using >90th percentile threshold) than among term AGA newborns in national datasets primarily from middle- and high-income settings (3). Others suggest no increased long-term mortality risk, except for a higher risk of death from specific conditions like digestive diseases and childhood malignancy (12,14,15)

Similarly, studies of the association between macrosomia and neonatal mortality remain inconsistent. An earlier analysis of 246,659 singleton term births from 373 facilities across 23 LMICs found that macrosomia (≥4000 g or ≥4500 g) was predictive of neonatal mortality and morbidity (16). However, in a recent descriptive multi-country analysis of 230,679 live births across 9 LMICs, there was no significant association between macrosomia (≥4000 g) and neonatal mortality (5). Hazel et al. also assessed fine strata of gestational age and birth weight; however, they did not investigate thresholds of LGA (>97th percentile) owing to an insufficient number of births, despite their potential to improve the identification of newborn vulnerability (17). These varied findings highlight the complexity of mortality risk assessment in LGA and macrosomic births, emphasizing the need for further research in LMICs, where data availability is low due to high out-of-health-facility births, inconsistent birth weight recording, and lack of reliable gestational age estimation.

To date, no study has assessed the specific contribution of LGA to neonatal mortality in LMICs, nor examined how this contribution varies across gestational age categories, including post-term births. This gap is particularly important given that post-term pregnancies are more common in LMICs (10–15%) than in high-income countries (2–5%), largely due to delayed antenatal care and limited access to labor induction (18).

Here, we addressed these gaps by quantifying the neonatal mortality rates of large newborns in LMIC cohorts between 2000 and 2015. Specifically, our objectives were to (1) estimate the mortality rates and relative risks for neonatal mortality for LGA (>90th and >97th percentiles overall, by gestational age categories and by term and post-term status) and for macrosomic (≥4000 g and ≥4500 g) newborns and (2) calculate the population attributable fraction (PAF) of neonatal mortality associated with LGA and macrosomia.

## METHODS

### Study design and data source

This study was a meta-analysis of individual participant data from subnational, population-based studies compiled for the Vulnerable Newborn Measurement Collaboration (Table S1). Dataset identification, data management, and cleaning have been described previously (19). In brief, subnational, population-based studies with high-quality birth outcome data were invited to join the Vulnerable Newborn Measurement Collaboration. To identify eligible datasets for inclusion, we conducted a comprehensive search of peer-reviewed databases (MEDLINE, Embase, Scopus, and OVID Global Health), clinical trial registries, and open data repositories (ClinicalTrials.gov, ISRCTN, Open Trials, and medRxiv). The search focused on terms such as “small for gestational age,” “low birthweight,” and “LMICs.”

Additionally, we created an online “call for data” survey, which was distributed through professional networks and investigator contacts. Principal investigators could submit their de-identified, individual-level datasets to the central analysis team or conduct analyses using standardized code and reporting templates.

Ethical approval for the primary studies was obtained from the respective ethics committees. The Vulnerable Newborn Measurement Collaboration also received ethical clearance from the Johns Hopkins Bloomberg School of Public Health (IRB No: 16439, approval date: 8 May 2021).

### Inclusion and exclusion criteria

Full study and individual-level inclusion and exclusion criteria are described elsewhere (19). In brief, all included studies reported information on birth weight, gestational age (assessed by ultrasound or last menstrual period), infant sex, and neonatal survival (19). Sixteen datasets met the predefined population-based recruitment criteria and included data on live births and deaths, high completeness (<30% missing values) for birth weight, gestational age, and sex. If 80% or more of births occurred in facilities, facility-based studies were considered population-based. Studies recruiting participants from antenatal clinics were deemed population-based if 90% of women in the sampled area had at least one antenatal care visit (4). Additionally, studies with birth weight missingness >70% among neonatal deaths were excluded. Two sets of studies (Burkina Faso 2004 and 2006 and Malawi 2003 and 2011) were combined into one Burkina Faso (2004 and 2006) and one Malawi (2003 and 2011) dataset since they originated from the same country and were collected by the same research teams, resulting in a total of 14 independent studies analyzed.

Individual records with improbable birth weights (<250 g or ≥6500 g), or birth weight for weekly gestational age greater than 5 standard deviations from the study level mean were excluded. Gestational age at delivery below 22 weeks or above 44 weeks were excluded owing to the lack of weights for these gestational ages in the extended version of INTERGROWTH-21st standards (4).

To estimate mortality risks, we assessed neonatal survival status at 28 days postdelivery, and infants lost to follow-up were excluded. Loss to follow up during the neonatal period was 5% or less in the included studies.

### Exposure and outcome definition

We assessed newborn size using the extended version of the INTERGROWTH-21st standards for size for gestational ages (20,21)

#### Exposure

Newborns were categorized by birth weight, gestational age, and size for gestational age (Box 1). As few births had a birth weight ≥5000 g, macrosomia analyses were restricted to the ≥4000 g and ≥4500 g thresholds

**Box 1:** Exposure definitions.

| Exposure | Categories |
| --- | --- |
| Birth weight | Reference: 2500–<4000 g; macrosomia grade 1: $\geq 4000$ g; grade 2: 4500–<5000 g |
| Gestational age | Preterm: <37 weeks; term: 37–<42 weeks (reference); post-term: $\geq 42$ weeks |
| Size for gestational age | AGA: 10th–90th percentile (reference); LGA: >90th percentile; >97th percentile where data permitted |

### Outcomes

Neonatal deaths were defined as newborns that died between 0 and 27 days after birth and further classified into early (0-6 days) and late (7-27 days) (3). Neonatal mortality risk was defined as the number of neonatal deaths per 1000 live births after improbable and missing exclusions.

### Statistical analysis

In many of these community-based trials, early neonatal deaths were missing birth weight, likely because the study team was unable to obtain a measurement before the death occurred. For this secondary analysis of mortality as an outcome, it was imperative to include these early neonatal deaths; therefore, we imputed birth weight for all missing values regardless of survival status. We used two approaches for birth weight imputation (5). In the first approach, we recalibrated all measured, non-missing birth weights to an estimated day 0. We then used these recalibrated birth weights to perform multiple imputation for missing values using a previously described model (5). This approach was applied to eight studies that measured birth weight throughout the early neonatal period (Table S1). In the second approach, four studies had no or very few birth weights measured after day 0, so recalibration would not improve the estimates (Table S1). For these studies, we used measured birth weights to directly impute missing values. Finally, in two studies, the proportion of missing birth weights was very low, making multiple imputation impractical; for these studies, we excluded birth weights measured >72 hours and used only measured birth weights. This imputation approach was applied only for the secondary data analysis; all primary studies included in the analysis have measured birth weights.

We used descriptive analysis (median and interquartile range [IQR]) to assess the median prevalence of LGA and macrosomic newborns and neonatal mortality rates among LGA and macrosomic newborns across the 14 studies. Neonatal mortality rates were summarized by preterm, term, or post-term category and size-for-gestational-age (preterm plus LGA, term plus LGA, post-term plus LGA) and macrosomia categories (≥4000 g and ≥4500 g) using medians and IQR. When no deaths occurred within a category despite a sufficient sample size, NMR was assigned 0 to allow for median and IQR calculations.

For each study, we estimated the relative risk, both unadjusted and adjusted for maternal education, age, parity, sex, and mode of delivery (when data were available and the model converged), with 95% confidence intervals, for large newborns using AGA as the reference group.

Unadjusted relative risk was defined as neonatal mortality rate in each exposure (LGA or macrosomia) group divided by the NMR in the reference group. We conducted a DerSimonian-Laird random-effects meta-analysis to generate pooled unadjusted relative risks with 95% confidence intervals. The Haldane correction was applied to handle zero-event cells, consistent with the approach used in comparable analyses in high-income country settings, facilitating direct comparison of estimates across contexts (3). The Haldane correction was used to mitigate zero or double-zero events (absence of events in both the target and control groups). Between-study heterogeneity was assessed by using the I² statistic. In addition, the Q statistic and corresponding *p*-value are also provided. Random- effects meta-analyses were conducted using both the imputed birth weights (main analysis) and the measured birth weights excluding the missing as a sensitivity analysis.

As a sensitivity analysis, and to address the zero-event issue in meta-analyses of binary outcomes, particularly when using relative risks as the effect size, we applied a beta-binomial regression model (22). This method is also considered for handling zero-event studies, especially when the meta-analysis includes few studies. We also considered a generalized linear mixed model. Due to convergence issues with the log-link, we used the logit link, which rather estimates the odds ratio; however, given that the outcome is rare, the odds ratio can be a reliable estimate of the relative risk.

We then performed adjusted relative risks to estimate the risk of neonatal mortality associated with LGA and macrosomia, accounting for residual confounding (maternal education, age, parity, sex, and mode of delivery). Generalized linear models with log-binomial regression were used to calculate adjusted relative risks. When no deaths occurred within a category despite a sufficient sample size, adjusted relative risks were not reported, owing to model convergence issues. The pooled adjusted relative risk estimates were derived by using a random-effects meta-analysis.

As a second sensitivity analysis, we also compared term AGA (reference group) with term LGA as previously done in their analysis of data from high-income countries (3).

Finally, the PAF for LGA and macrosomia was calculated with the following formula, where *Pe* represents the prevalence of each large newborn type and *RR* the meta-analytic relative risk of mortality associated with each large newborn type (LGA >90th percentile and LGA >97th percentile) and macrosomia (≥4000 g):

Equation 1:

The global prevalences of LGA >90th percentile and macrosomia ≥4000 g and ≥4500 g were derived from our recent publication, which included 45 studies from 23 countries comprising 476,939 live births (9) whereas the prevalence of LGA >97th percentile is reported from the current study. The variance of PAF was estimated by using the delta method (23) and the corresponding 95% CI was then calculated.

All analyses, except for the beta binomial regression models (SAS version 9.4) were conducted in R version 4.4.3 (R Foundation for Statistical Computing, Vienna, Austria).

## RESULTS

Of the 14 datasets from the 16 prospective cohort studies conducted between 2000 and 2017 across 9 LMICs included in the analysis, 6 were from Asia, 7 from Africa, and 1 from the Americas. These studies included 134,952 live births with data on neonatal mortality and birth outcomes.

### Descriptive analysis results

#### Prevalence of LGA and macrosomia

The prevalence of LGA varied substantially across sites and ranged from 0.9% to 22.1% for the >90th percentile and 0.3% to 15.1% for the>97th percentile (Table 1). Overall, the median LGA (>90th percentile) and median LGA (>97th percentile) prevalence was 5.3% (IQR 3.6, 8.2) and 2.6% (IQR 1.3, 4.5), respectively. The median macrosomia (≥4000 g) and (≥4500 g) prevalences accounted for 1.0% (IQR 0.3, 3.1) and 0.06% (IQR 0.01, 0.30) of live births, respectively, and ranged from 0.1% to 5.0% for ≥4000 g and 0.0% to 0.6% for ≥4500 g. Overall, 21 infants across all cohorts weighed ≥5000 g (Table 2).

**Table 1.**
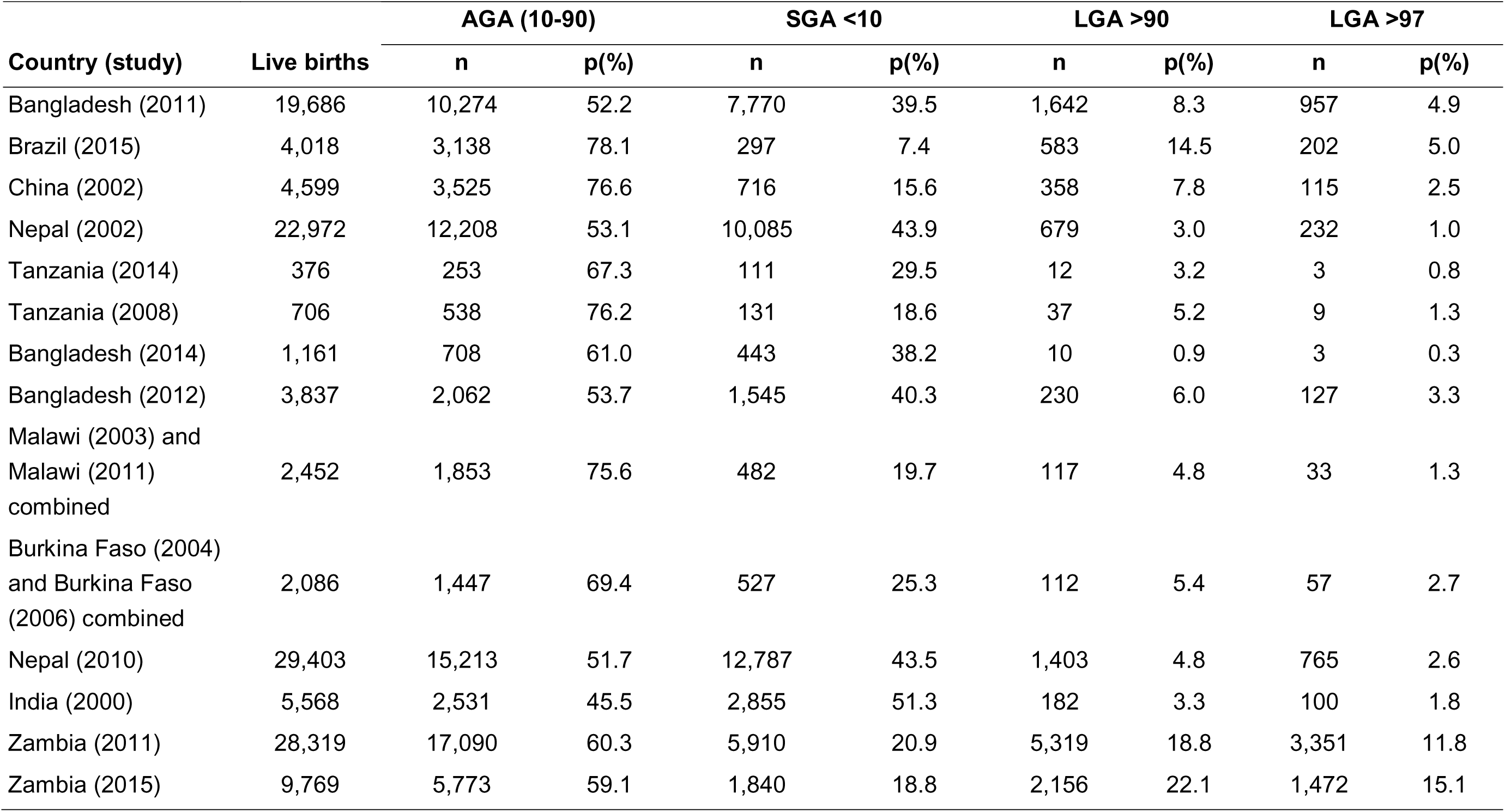
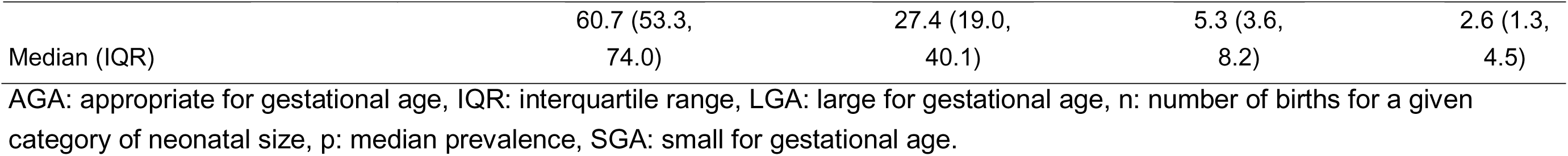
Prevalence of LGA >90th centile, LGA>97th centile in studies included in the individual participant data meta-analysis.

**Table 2.**
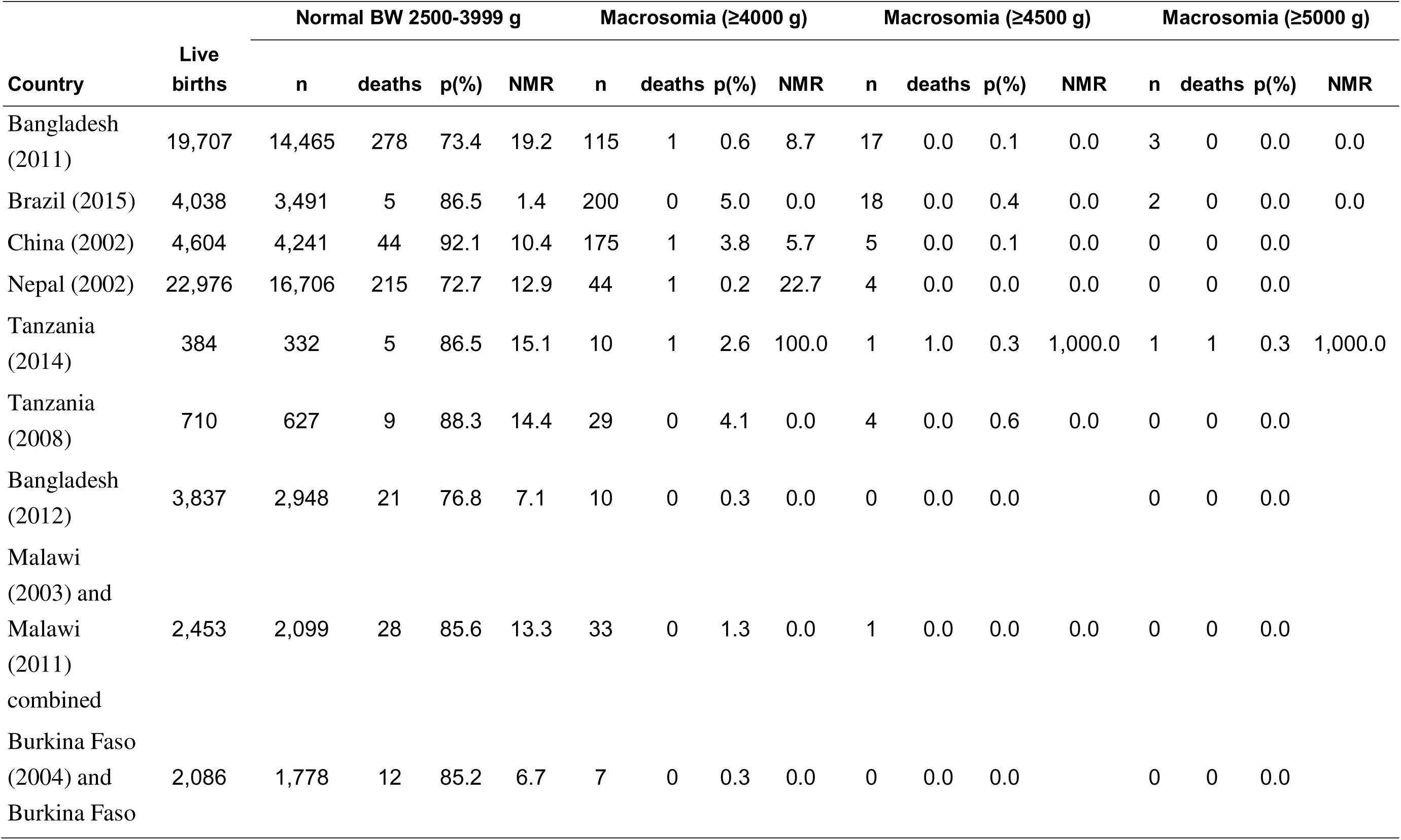
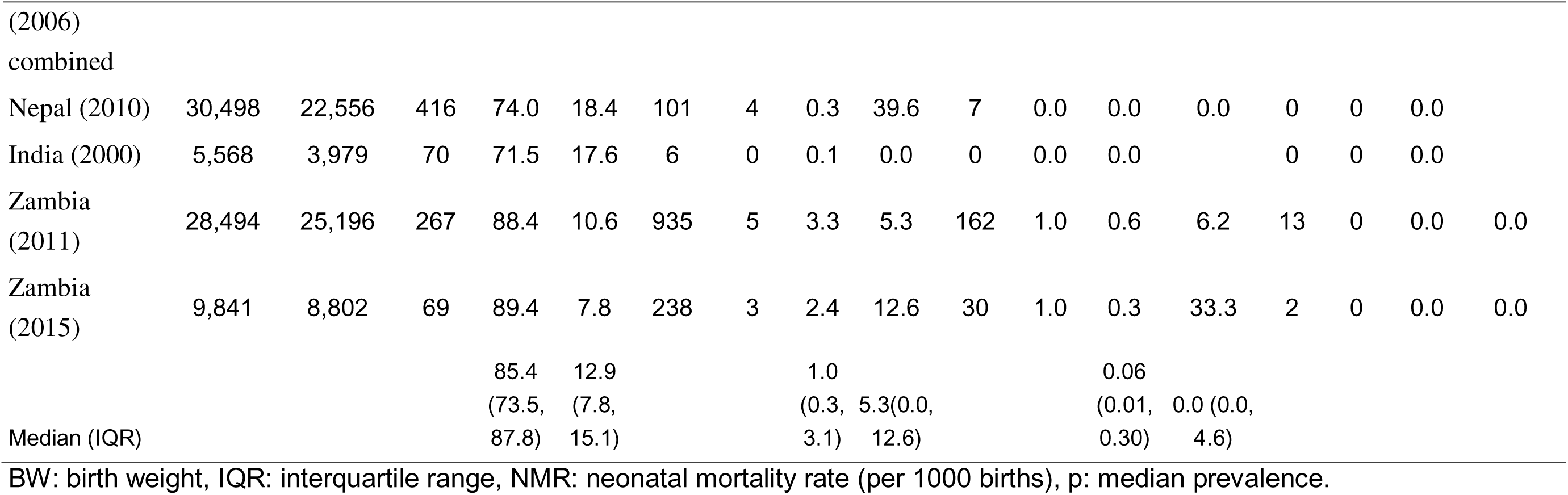
Combined neonatal NMR of macrosomic neonates (≥4000 g; ≥4500 g and ≥5000 g).

#### Neonatal mortality rate of LGA and macrosomia

Neonatal mortality rate was highest among preterm LGA infants (median, 61.3 deaths per 1000 live births; IQR 13.8, 92.5). Post-term LGA births were rare, and no deaths were reported in most sites (Table 3). Neonatal mortality among macrosomic infants was generally low (median 5.3 per 1000 live births; IQR 0.0, 12.6) and remained low for ≥4500 g (median 0.0; IQR 0.0, 4.6 Table 2).

**Table 3.**
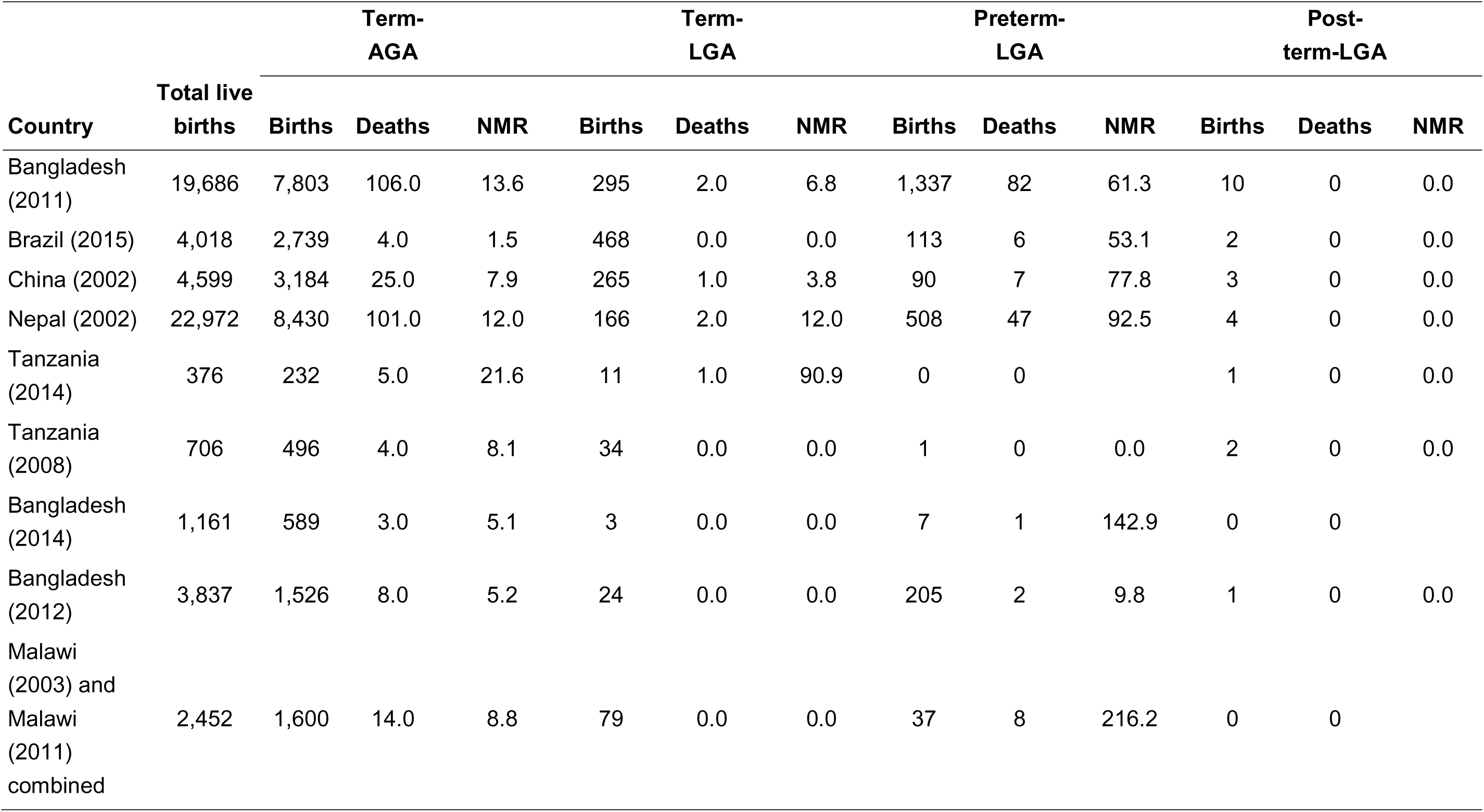
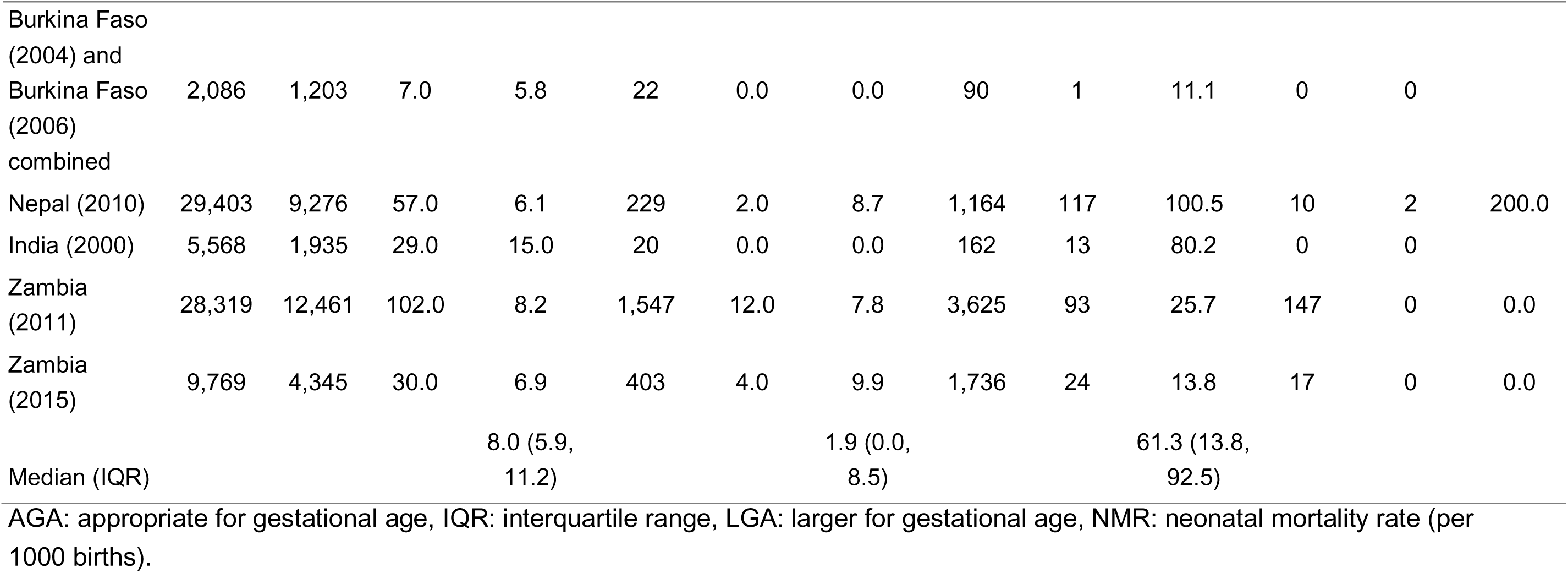
Neonatal mortality rate of LGA (>90 centile) neonates by cohort and term.

| Country | Total live births | Term-AGA |  |  | Term-LGA |  |  | Preterm-LGA |  |  | Post-term-LGA |  |  |
| --- | --- | --- | --- | --- | --- | --- | --- | --- | --- | --- | --- | --- | --- |
|  |  | Births | Deaths | NMR | Births | Deaths | NMR | Births | Deaths | NMR | Births | Deaths | NMR |
| Bangladesh (2011) | 19,686 | 7,803 | 106.0 | 13.6 | 295 | 2.0 | 6.8 | 1,337 | 82 | 61.3 | 10 | 0 | 0.0 |
| Brazil (2015) | 4,018 | 2,739 | 4.0 | 1.5 | 468 | 0.0 | 0.0 | 113 | 6 | 53.1 | 2 | 0 | 0.0 |
| China (2002) | 4,599 | 3,184 | 25.0 | 7.9 | 265 | 1.0 | 3.8 | 90 | 7 | 77.8 | 3 | 0 | 0.0 |
| Nepal (2002) | 22,972 | 8,430 | 101.0 | 12.0 | 166 | 2.0 | 12.0 | 508 | 47 | 92.5 | 4 | 0 | 0.0 |
| Tanzania (2014) | 376 | 232 | 5.0 | 21.6 | 11 | 1.0 | 90.9 | 0 | 0 |  | 1 | 0 | 0.0 |
| Tanzania (2008) | 706 | 496 | 4.0 | 8.1 | 34 | 0.0 | 0.0 | 1 | 0 | 0.0 | 2 | 0 | 0.0 |
| Bangladesh (2014) | 1,161 | 589 | 3.0 | 5.1 | 3 | 0.0 | 0.0 | 7 | 1 | 142.9 | 0 | 0 |  |
| Bangladesh (2012) | 3,837 | 1,526 | 8.0 | 5.2 | 24 | 0.0 | 0.0 | 205 | 2 | 9.8 | 1 | 0 | 0.0 |
| Malawi (2003) and Malawi (2011) combined | 2,452 | 1,600 | 14.0 | 8.8 | 79 | 0.0 | 0.0 | 37 | 8 | 216.2 | 0 | 0 |  |
| Burkina Faso (2004) and Burkina Faso (2006) combined | 2,086 | 1,203 | 7.0 | 5.8 | 22 | 0.0 | 0.0 | 90 | 1 | 11.1 | 0 | 0 |  |
| Nepal (2010) | 29,403 | 9,276 | 57.0 | 6.1 | 229 | 2.0 | 8.7 | 1,164 | 117 | 100.5 | 10 | 2 | 200.0 |
| India (2000) | 5,568 | 1,935 | 29.0 | 15.0 | 20 | 0.0 | 0.0 | 162 | 13 | 80.2 | 0 | 0 |  |
| Zambia (2011) | 28,319 | 12,461 | 102.0 | 8.2 | 1,547 | 12.0 | 7.8 | 3,625 | 93 | 25.7 | 147 | 0 | 0.0 |
| Zambia (2015) | 9,769 | 4,345 | 30.0 | 6.9 | 403 | 4.0 | 9.9 | 1,736 | 24 | 13.8 | 17 | 0 | 0.0 |
| Median (IQR) |  |  |  | 8.0 (5.9, 11.2) |  |  | 1.9 (0.0, 8.5) |  |  | 61.3 (13.8, 92.5) |  |  |  |

AGA: appropriate for gestational age, IQR: interquartile range, LGA: larger for gestational age, NMR: neonatal mortality rate (per 1000 births).

## Results of meta-analysis

### Neonatal mortality risk associated with LGA

The pooled risk ratio for LGA >90th percentile was 2.46 (95% CI: 1.86–3.25) (Figure 1A), while the pooled relative risk for LGA >97th percentile was 3.77 (95% CI: 2.50–5.69) (Figure 1B). Using a subset of studies with available data, we calculated the adjusted relative risk for LGA at both the 90th and 97th percentile thresholds. A similar pattern emerged, with the 97th percentile threshold (RR 2.70; 95% CI: 1.14–6.38) demonstrating a higher mortality risk compared to the 90th percentile threshold (RR 2.33; 95% CI: 1.63–3.33) (Figures 2A and B). The adjusted relative risks were attenuated compared to the crude estimates, likely due to confounding by the factors controlled for in the adjusted models. However, because the adjusted relative risk analysis was conducted on a subset of studies, direct comparisons between the crude and adjusted estimates should be interpreted with caution.

**Figure 1.**
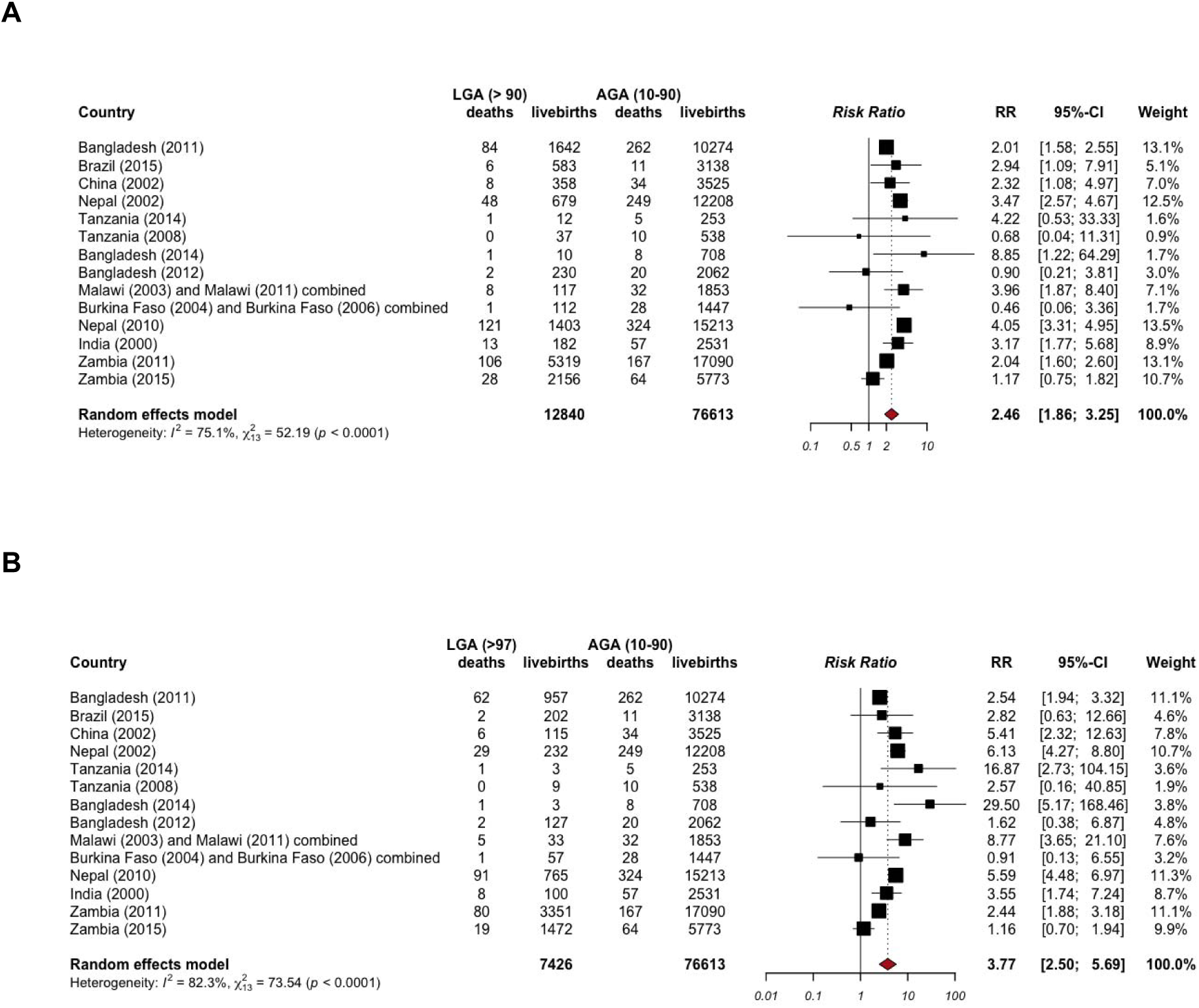
Pooled mortality risk of **(A)** LGA >90th percentile neonates and **(B)** LGA >97th percentile neonates. AGA, appropriate for gestational age; LGA, large for gestational age; RR, relative risk. All pooled estimates were derived from imputed data.

**Figure 2.**
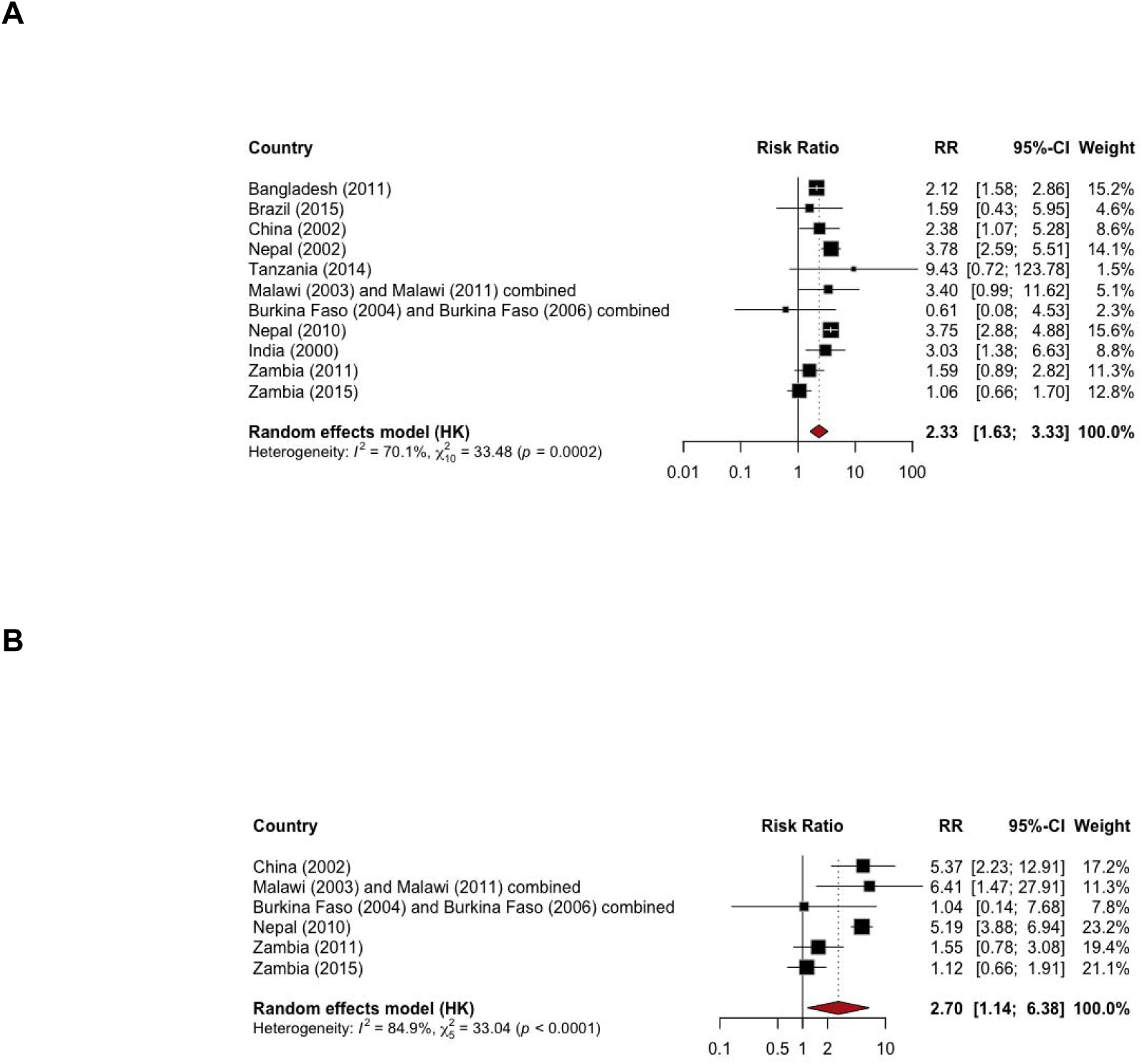
Pooled adjusted mortality risk of **(A)** LGA >90th percentile and **(B)** LGA >97th percentile. RR, relative risk.

Most of the deaths were concentrated in the preterm LGA category. When we examined term plus LGA only, we found fewer deaths and only elevated mortality risk for the 97th threshold (3.14; 95% CI: 1.58-6.22) but not the 90th threshold (1.14; 95% CI: 0.78-1.68) (Figures 3A and B). Overall, the neonatal mortality risk among post-term LGA infants (>90th percentile) was 10.2 per 1000 births (2/197), similar with 10.2 per 1000 births (65/6,348) among post-term AGA infants, with 7 of the 9 studies reporting fewer than 10 births in the LGA group. The neonatal mortality risk among post-term LGA infants (>97th percentile) was 0 (0/72).

**Figure 3.**
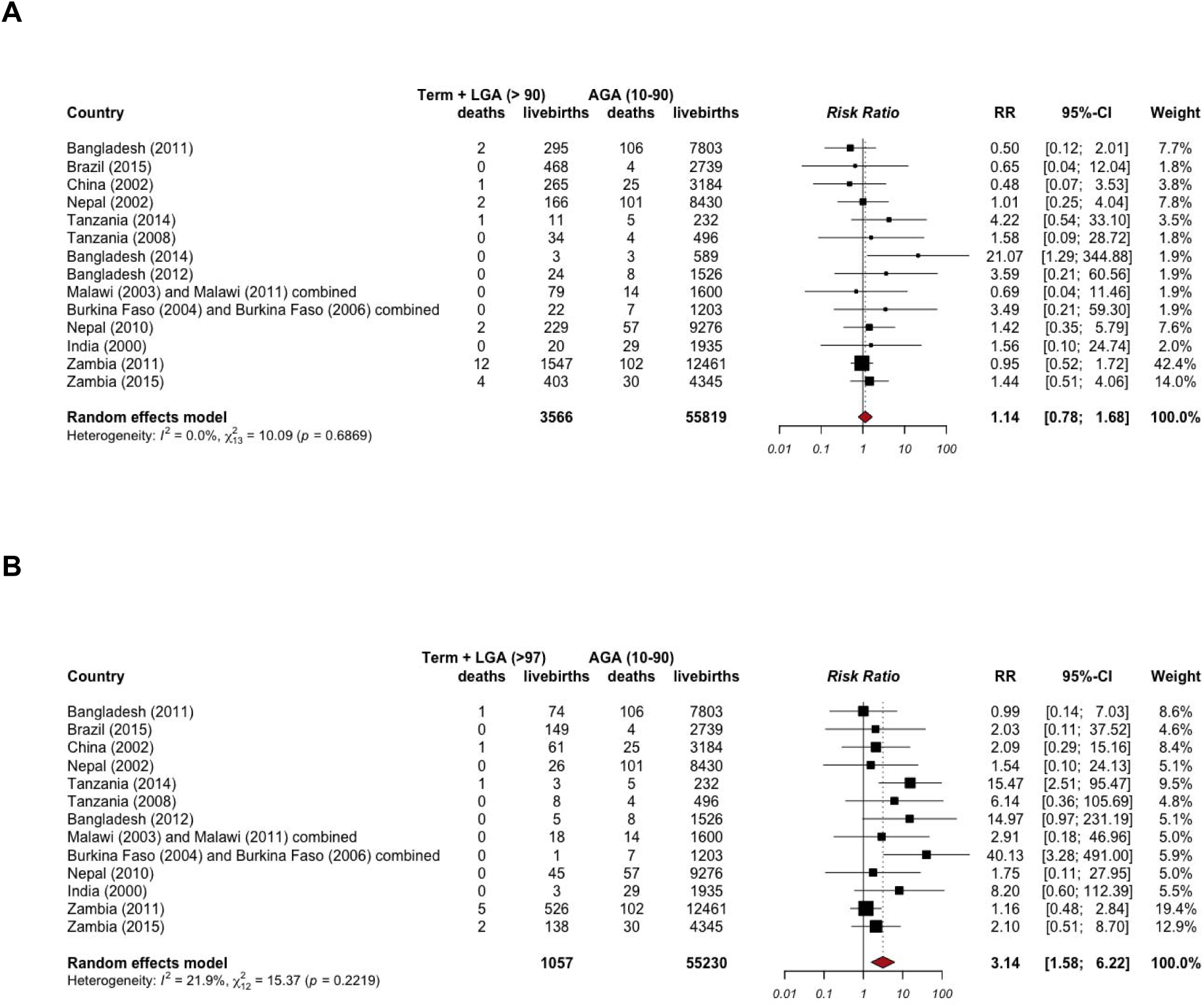
Pooled mortality risk of **(A)** term+LGA >90th percentile neonates and **(B)** term+LGA >97th percentile neonates compared to term AGA. AGA, appropriate for gestational age; LGA, large for gestational age; RR, relative risk. All pooled estimates were derived from imputed data.

Stratified analyses showed consistently elevated risks in both early and late neonatal periods. Among those in the LGA >90th percentile, pooled relative risks were 2.71 (95% CI: 1.92-3.82) for early neonatal mortality and 1.69 (95% CI: 1.22-2.34) for late mortality (Figures 4A and B). Risks were even greater among those in the LGA >97th percentile. Relative risk was 4.46 (95% CI: 2.75-7.22) for early neonatal mortality and 2.25 (95% CI: 1.43-3.52) for late neonatal mortality (Figures 4C and D)

**Figure 4.**
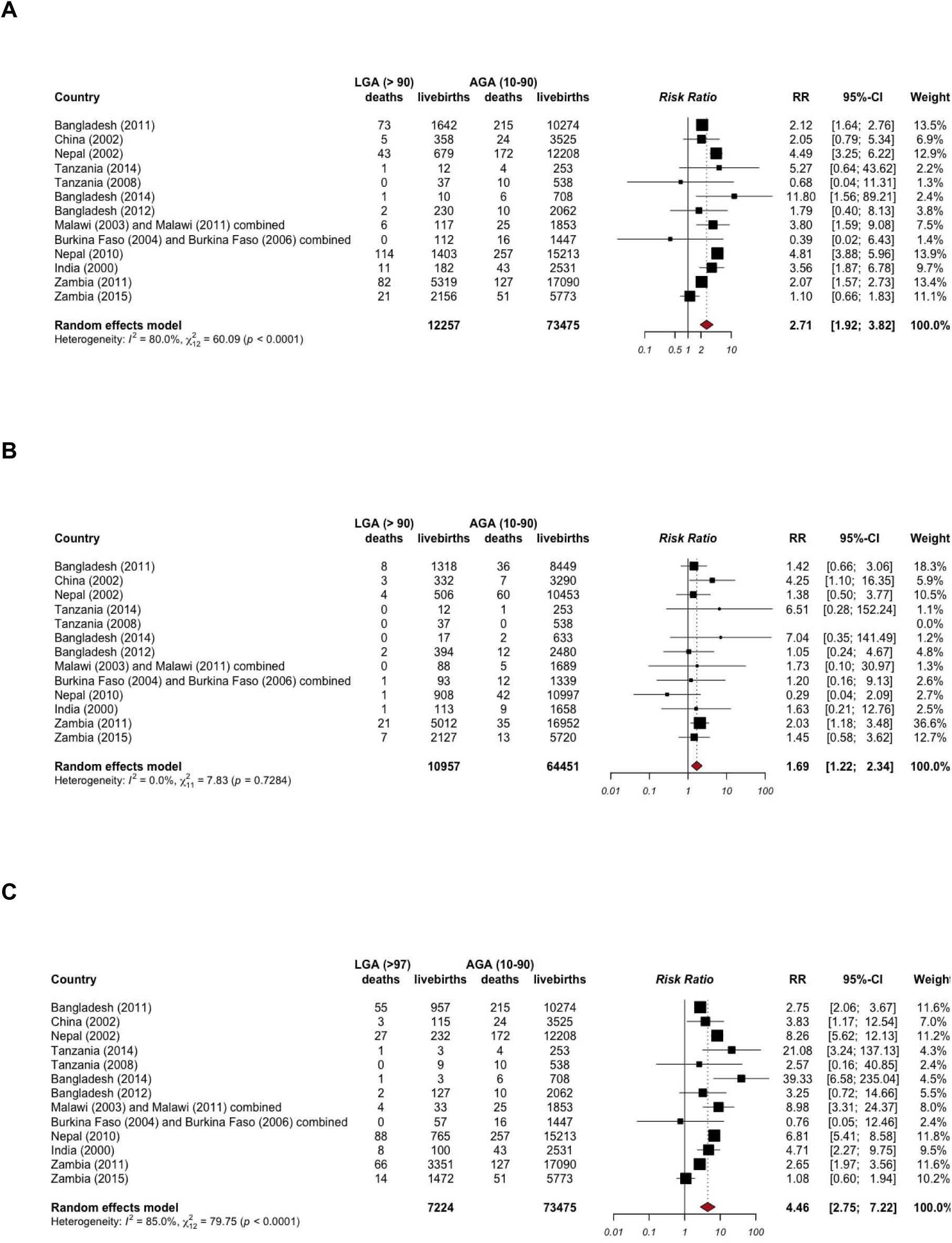
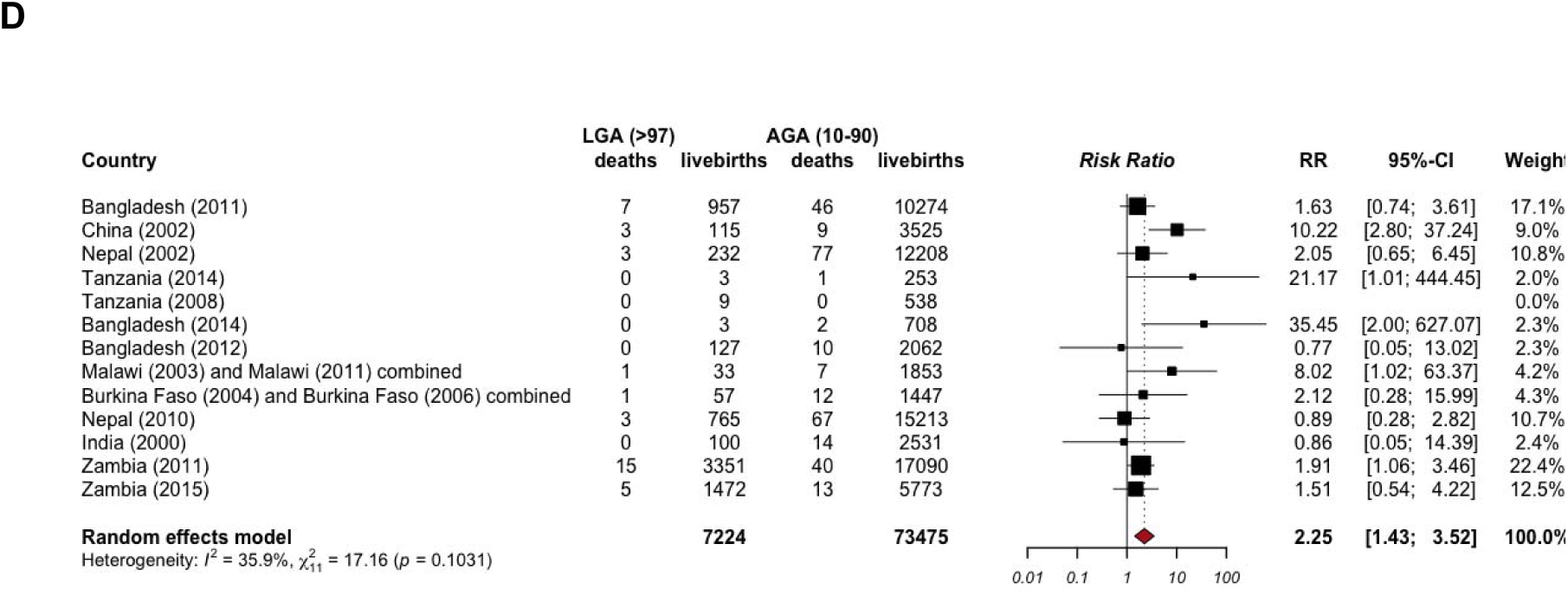
**(A)** Pooled early mortality risk of LGA >90th percentile neonates. **(B)** Pooled late mortality risk of LGA >90th percentile neonates. **(C)** Pooled early mortality risk of LGA >97th percentile neonates. **(D)** Pooled late mortality risk of LGA >97th percentile neonates. AGA, appropriate for gestational age; LGA, large for gestational age; RR, relative risk. All pooled estimates were derived from imputed data.

Finally, we replicated this analysis using only measured birth weights, excluding cases with missing data, and observed a similar pattern with one notable exception (Figures S1–S11). Compared to the risk estimates from the calibrated data, the mortality risk during the early neonatal period (RR 1.47; 95% CI: 1.16–1.86) was lower than during the late neonatal period (RR 1.69; 95% CI: 1.22–2.34

### Neonatal mortality risk associated with macrosomia

Our analysis showed very few macrosomia-related infant deaths; there was no association between macrosomia (≥4000 g) and neonatal mortality risk (RR 1.44; 95% CI: 0.82-2.55) compared with infants weighing 2500–<4000 g (Figure 5).

**Figure 5.**
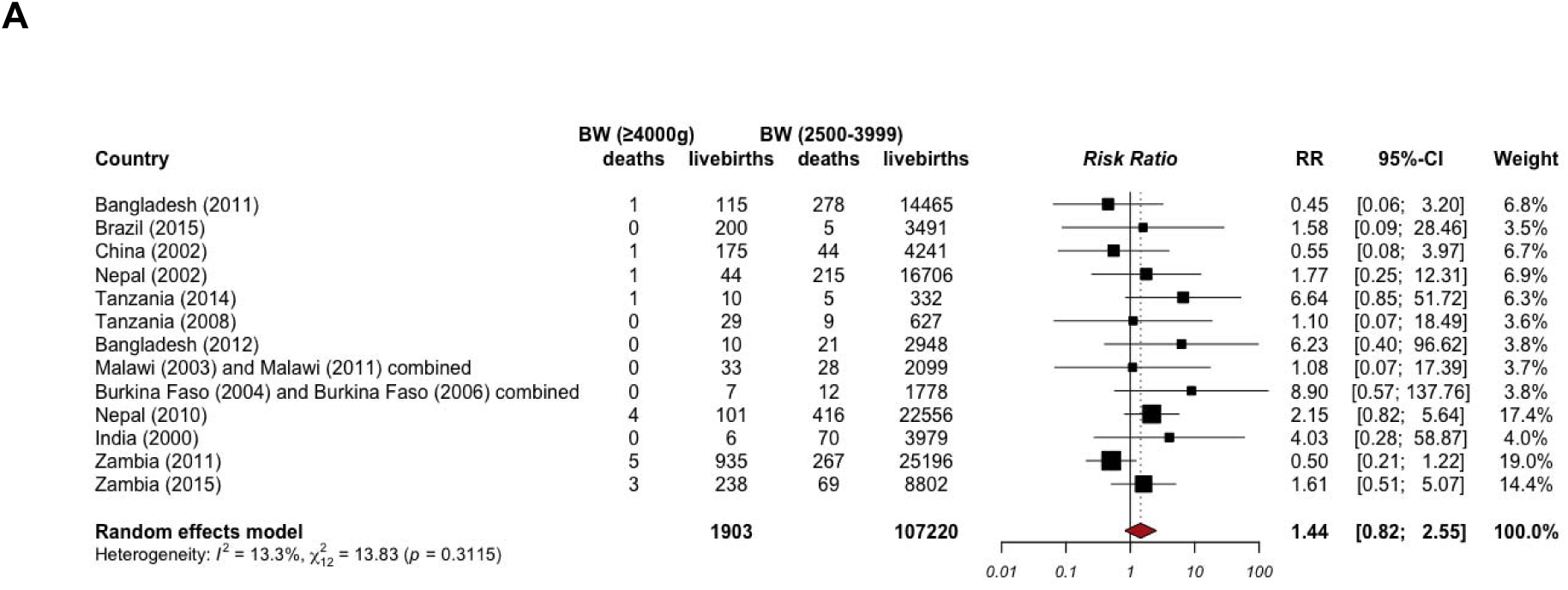
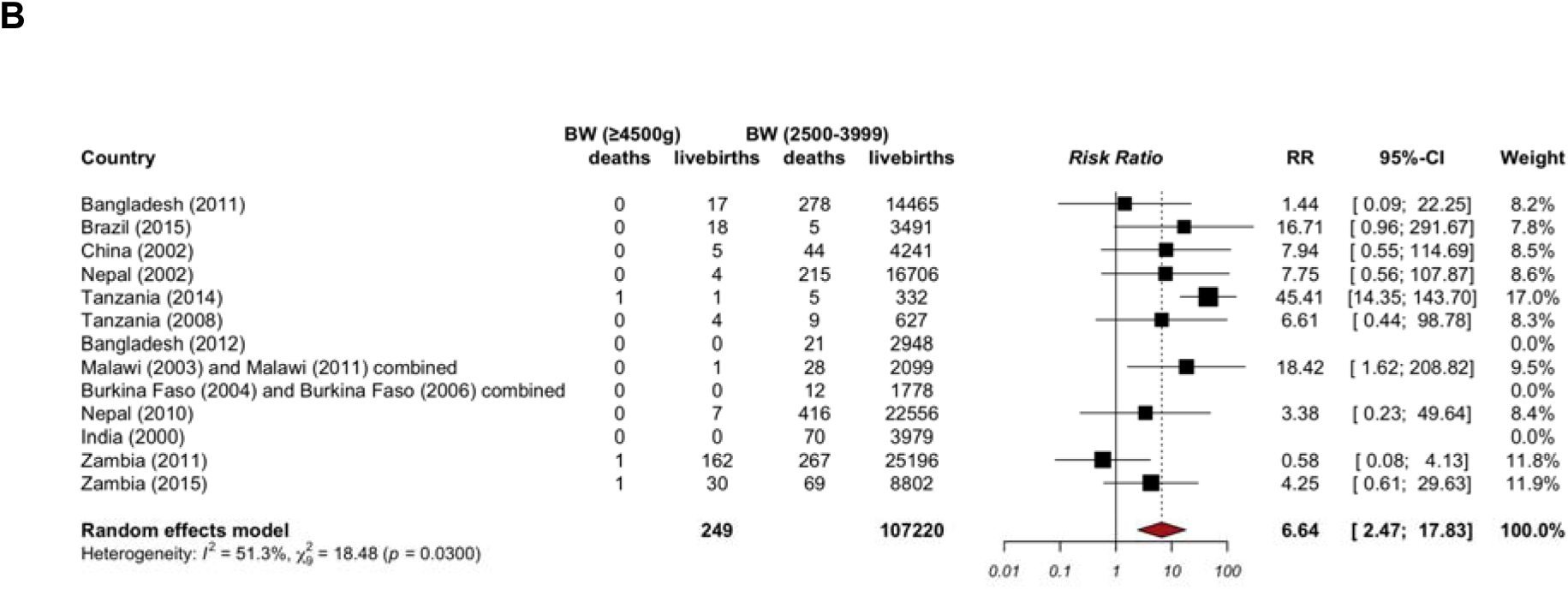
**(A)** Pooled relative risks (RRs) between macrosomia ≥4000 g and neonatal mortality (0-27 days) **(B).** Pooled relative risks (RRs) between macrosomia ≥4500 g and neonatal mortality (0-27 days). BW, birth weight. All pooled estimates were derived from imputed data.

Stratified analysis by neonatal period showed an elevated mortality risk (RR 3.15; 95% CI: 1.59-6.25) in the late neonatal period for macrosomic (≥4000 g) neonates compared with those with normal birth weight. There was no association in the early neonatal period (Figure 6).

**Figure 6.**
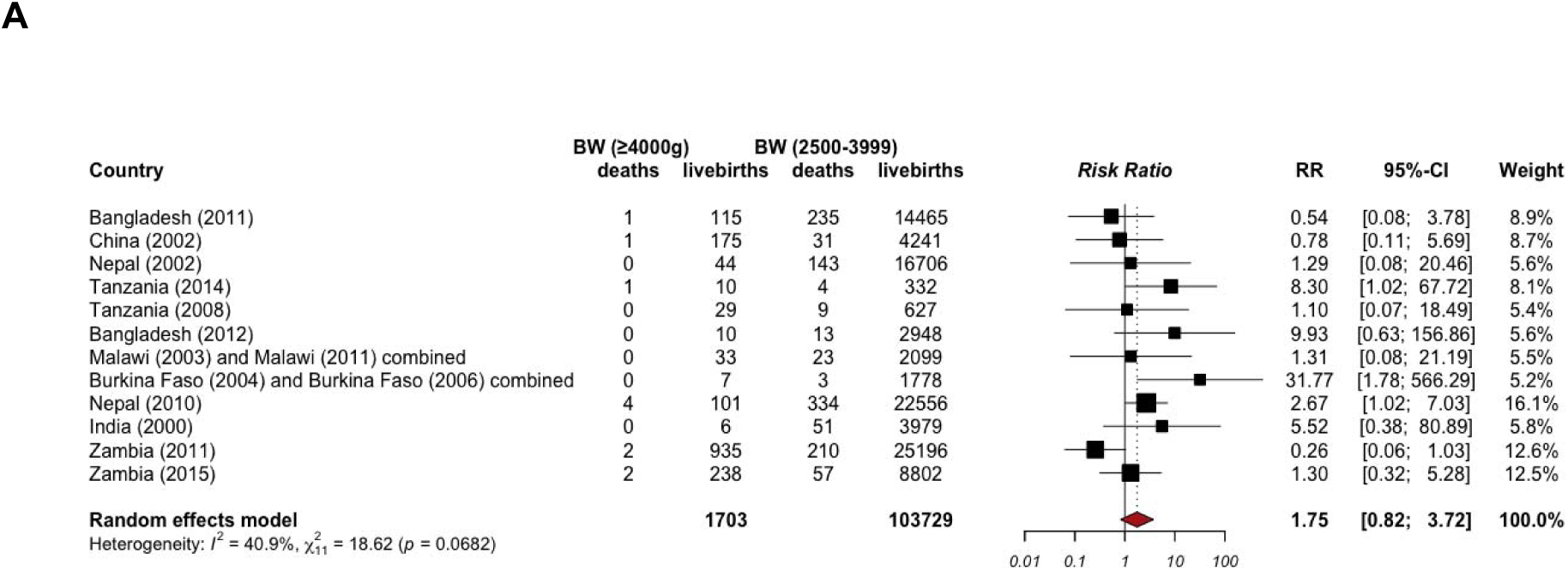
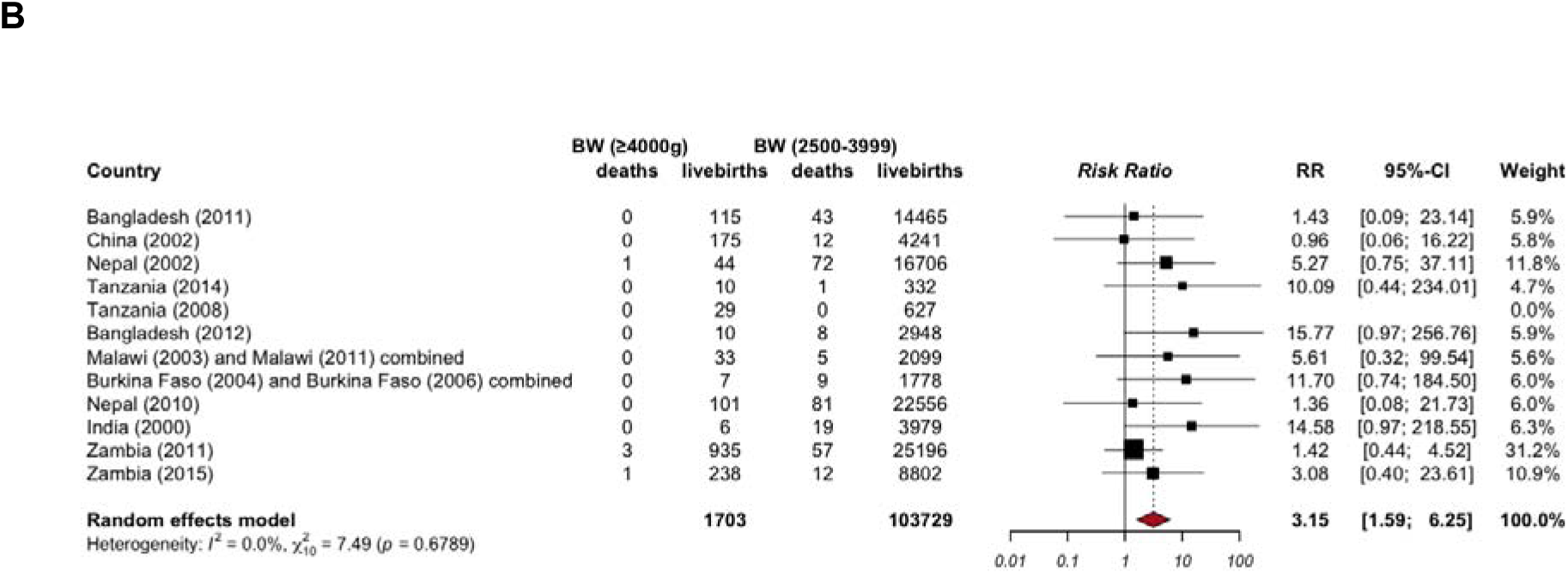
**(A)** Pooled relative risks (RRs) between macrosomia ≥4000g and early (0-6 days) neonatal mortality. **(B)** Pooled RRs between macrosomia ≥4000g and late (7-28 days) neonatal mortality. BW, birth weight. All pooled estimates were derived from imputed data.

Sensitivity analyses using alternative modeling approaches yielded attenuated and statistically nonsignificant associations. Using the generalized linear mixed model, macrosomia was not associated with neonatal mortality overall (OR = 0.80; 95% CI: 0.49–1.32), nor with early (OR = 0.72; 95% CI: 0.39–1.31) or late neonatal mortality (OR = 1.15; 95% CI: 0.47–2.80). Similarly, beta-binomial regression models showed no evidence of an increased risk of neonatal mortality among macrosomia infants overall (RR = 0.86; 95% CI: 0.45–1.65), or when considering early (RR = 0.83; 95% CI: 0.40–1.71) and late neonatal mortality (RR = 0.97; 95% CI: 0.38–2.49) (Tables S2 and S3).

Overall, the neonatal mortality risk among macrosomia infants (≥4500) was 12.1 per 1000 births (3/249), compared with 13.4 per 1000 births (1439/107,220) among infants weighing 2500-<4000 g, with 10 of the 14 studies reporting fewer than 10 births in the LGA group.

Although the mortality risk (RR 6.64; 95% CI: 2.47-17.83; Figure 4B) was elevated for macrosomia ≥ 4500 g compared with normal birth weight in the main analysis, sensitivity analyses using other modeling strategies (generalized linear mixed model and beta-binomial models) showed no association (RR = 1.15; 95% CI: 0.36–3.66; Tables S2 and S3).

### PAF analyses

Overall, the population attributable fraction (PAF) associated with LGA >90th percentile was 7.2% (95% CI: 4.1%–10.3%) of neonatal deaths in the target population. The corresponding PAFs were 8.3% (95% CI: 4.2%–12.4%) for early neonatal deaths and 3.2% (95% CI: 0.9%–5.5%) for late neonatal deaths. Similarly, LGA >97th percentile accounted for 6.7% (95% CI: 3.2%–10.2%) of neonatal deaths in the target population, with PAFs of 8.3% (95% CI: 3.5%–13.0%) for early and 3.2% (95% CI: 0.7%–5.7%) for late neonatal deaths. For macrosomia, defined as birth weight ≥4000 g, the overall PAF was 0.4% (95% CI: 0.0%–1.2%). The PAFs for macrosomia ≥4000 g were 0.7% (95% CI: 0.0%–2.0%) for early neonatal deaths and 2.1% (95% CI: 0.0%–4.2%) for late neonatal deaths (Table 4).

**Table 4.** Population attributable fractions (PAFs) for neonatal mortality associated with large-for-gestational-age (LGA)

| Main analyses | p% (IQR) | N | PAF% (95% CI) |  |  |
| --- | --- | --- | --- | --- | --- |
|  |  |  | Overall | Early | Late |
| LGA90 | 5.3 (3.6, 8.2) | 476,939 | 7.2 (4.1-10.3) | 8.3 (4.2-12.4) | 3.2 (0.9-5.5) |
| LGA97 | 2.6 (1.3, 4.5) | 134,952 | 6.7 (3.2-10.2) | 8.3 (3.5-13.0) | 3.2 (0.7-5.8) |
| Macrosomia > 4000g | 0.06 (0.0, 0.30) | 313064 | 0.4 (0.0-1.2) | 0.7 (0.0-2.0) | 2.1(0.0-4.2) |

CI, confidence interval; IQR, interquartile range; LGA90, large-for-gestational-age >90th centile; LGA97, large-for-gestational-age >97th centile; p, median prevalence; PAF, population attributable fraction.

## DISCUSSION

In this multi-country analysis of 134,952 live births from 16 LMIC birth cohorts, we observed elevated median neonatal mortality risks, with preterm LGA infants experiencing the highest overall median neonatal mortality rate. Both LGA and macrosomia infants had higher neonatal mortality than AGA or normal-birth weight infants. However, associations for macrosomia were less consistent across analyses. LGA infants above the 90th percentile had more than a twofold higher risk of neonatal death, while those above the 97th percentile had an approximately threefold higher risk, with similar patterns observed when compared specifically with term AGA infants. For both LGA thresholds, excess mortality risk was more pronounced in the early neonatal period than in the late neonatal period. Population-attributable fraction analyses indicated that a modest but non-negligible proportion of neonatal deaths could be attributed to LGA and macrosomia, with slightly higher fractions observed for LGA in the early neonatal period.

Evidence from LMICs on the risks associated with LGA and macrosomia has been limited. A facility-based, multi-country study, using data from WHO Global Survey on Maternal and Perinatal Health to examine births in 23 LMICs, found that increased perinatal mortality was associated with macrosomia, particularly in Africa, but not in Latin America or Asia, Similarly, a recent study from Bangladesh reported that the mortality rate was higher among LGA infants than among term AGA infants (24). In contrast, a more recent descriptive multi-country analysis of 230,679 live births across 9 LMICs reported no significant association between macrosomia defined as ≥4000 g and neonatal mortality; however, that study did not assess higher-risk thresholds, such as LGA above the 97th percentile (5). Our study adds to this evidence by using individual-level data across diverse LMIC cohorts. We demonstrate a stepwise increase in neonatal mortality risk, particularly among infants above the 97th percentile. Moreover, although neonatal death was rare among infants weighing ≥4500 g, the point estimate was consistent with an increased mortality risk in this group, suggesting, excess neonatal mortality.

These findings are inconsistent with evidence from high-income countries, where LGA and macrosomia have been linked to increased perinatal mortality, but typically only at birth weights ≥4500 g, not ≥4000 g (3). The lower absolute risk observed among large infants in HICs likely reflects better access to intrapartum monitoring, timely caesarean delivery, and advanced neonatal care. The risk associated with birth weight ≥4000 g may also vary across settings due to differences in maternal size and population birth weight distributions. In LMICs, where women are generally smaller, birth weights above 4000 g may confer higher perinatal and maternal risks, whereas in HICs, larger maternal size may mitigate some of these risks. This aligns with evidence that macrosomia adverse outcomes are population-specific and influenced by maternal anthropometry (25–27).

Notably, the highest median neonatal mortality rate was observed among preterm LGA infants, highlighting the profound biological and clinical vulnerability of this subgroup. This finding likely reflects a compounded vulnerability, whereby these fetuses are simultaneously exposed to the physiological immaturity of prematurity and the metabolic dysregulation associated with excessive fetal growth. In many LMIC settings, where universal screening for gestational diabetes remains limited, LGA status may function as a clinical proxy for undiagnosed maternal hyperglycemia and consequent fetal hyperinsulinemia (27)

From a pathophysiological perspective, fetal hyperinsulinism has been shown to impair pulmonary maturation by inhibiting surfactant synthesis and disrupting glucocorticoid-mediated differentiation of type II pneumocytes (28–30). As a result, despite their increased somatic growth, these infants may exhibit functionally immature lungs, predisposing them to respiratory distress syndrome and neonatal mortality.

Additionally, our stratified results suggest that the elevated risks associated with LGA and macrosomia extend beyond the early neonatal period into the late neonatal phase. This finding aligns with previous research indicating that complications among large infants, such as feeding difficulties, hypoglycemia, and undiagnosed infections, can persist beyond the immediate intrapartum period (31,32). This pattern likely reflects the concentration of severe perinatal complications in the first days of life, including birth asphyxia, traumatic delivery, respiratory compromise, and metabolic instability, which disproportionately affect large infants and are most lethal shortly after birth. In settings with limited access to timely obstetric and neonatal care, delays in recognition and management of these complications may further amplify early neonatal mortality (33).

To assess the robustness of these findings, we repeated the analysis using only measured birth weights and excluding cases with missing data, yielding largely consistent results with the main analysis with one notable exception: The risk associated with LGA appeared lower during the early neonatal period than in the late neonatal period. This discrepancy is likely explained by the higher likelihood of missing birth weight data among early neonatal deaths, which may bias estimates when such cases are excluded. These findings underscore the critical importance of accounting for missing data, particularly through birth weight imputation, in neonatal mortality analyses.

Another important observation is the substantial heterogeneity in risk estimates across study sites. This variation may reflect differences in underlying maternal risk factors, including undiagnosed gestational diabetes, nutritional exposures, obesity prevalence, and disparities in the availability and quality of obstetric and neonatal care. These findings underline the context-specific interplay between maternal health, health system capacity, and neonatal outcomes.

### Interpretation and implications

The elevated mortality risk among LGA infants highlights the urgent need to expand neonatal survival strategies in LMICs. Antenatal interventions, such as screening for gestational diabetes, nutritional counseling, and maternal weight monitoring remain underutilized but have the potential to reduce the incidence and complications of excessively large newborns (34–38). This gap is particularly concerning given the rapidly increasing prevalence of diabetes in LMICs, where GDM is also on the rise (39). Yet these interventions hold significant potential to reduce both the incidence of excessive fetal growth and the associated complications among large-for-gestational-age newborns. In settings where post-term pregnancies are common and access to labor induction is limited, our findings suggest that improved assessment of fetal growth and timely consideration of delivery planning, within existing guideline frameworks, may be relevant for mitigating neonatal risks associated with LGA, alongside strengthened intrapartum and neonatal care (38).

Given that neonatal mortality constitutes an increasing proportion of deaths among children in LMICs (40–42), failing to address risks at both ends of the birth weight spectrum may hinder progress toward United Nations Sustainable Development Goal targets (43). Our findings reinforce the need for a “dual burden” approach that integrates vulnerabilities associated with both small and large newborns into neonatal survival strategies. Global agendas on newborn survival have traditionally centered on low birth weight and preterm infants, given their well-documented contribution to neonatal mortality (2,44–46). However, LGA infants also face elevated risks, and a measurable share of neonatal deaths is linked to these high-risk infants, which, if neglected, will continue to contribute to preventable neonatal deaths in LMICs.

### Strengths and limitations

Key strengths of this study include the large, multi-country sample and the use of harmonized individual participant data for both LGA and macrosomia, which enabled consistent definitions and robust adjustment for potential confounders. Stratification by gestational age and neonatal period enhanced the granularity of risk assessment, and findings remained consistent across multiple sensitivity analyses with imputed data. In addition, we complemented continuity correction–based meta-analyses with alternative approaches, including beta-binomial models and generalized linear mixed-effects models, which have been shown to perform better in the presence of rare events, thereby strengthening the robustness of our findings.

However, several limitations should be acknowledged. First, the data included in this analysis were collected between 2000 and 2017, and maternal, obstetric, and neonatal care practices, as well as clinical guidelines, may have changed since then, potentially affecting the generalizability of our findings to current settings. Variability in the measurement and recording of gestational age across study sites may also have affected the classification of LGA status. Despite adjustment for known confounders, residual confounding cannot be ruled out, particularly regarding maternal diabetes, which was inconsistently measured across cohorts. A further limitation is that although evaluating the contribution of LGA in post-term births and macrosomia among infants weighing ≥4500 g was one of our objectives, we were unable to perform robust analyses due to the small sample size in these subgroups. In addition, while the primary meta-analysis suggested an increased risk of neonatal mortality among macrosomia infants, this finding was not consistent in sensitivity analyses using alternative modeling approaches, reflecting substantial uncertainty. Larger studies or pooled multi-country data are needed to better characterize these risks and guide timely interventions, particularly as post-term fetal overgrowth combined with prolonged gestation may further increase neonatal mortality.

An additional limitation concerns the heterogeneity in gestational age assessment methods across the included studies. Several studies used different approaches, with some combining multiple methods for gestational age estimation, potentially affecting the consistency and comparability of the findings. Furthermore, the limited number of studies in certain subgroups or specific contexts constrained further exploratory analyses and limited the ability to draw robust conclusions.

Substantial heterogeneity was observed across study sites (I² = 75.1% and 82.3% for LGA at the 90th and 97th percentiles, respectively), likely reflecting contextual differences in maternal risk factors, clinical practices, and health system capacities. Data on the prevalence of LGA and macrosomia used to compute the PAF were available for only some countries, and in several instances the data were either outdated or not representative of the entire population. Having incomplete data may limit the accuracy of our estimates in reflecting the true global burden of neonatal mortality associated to LGA and macrosomia. Additional high-quality, population-based studies are needed in underrepresented regions to strengthen and validate these findings.

Also, while we considered exploring differences between low-income and middle-income countries, as well as between continents, the limited number of studies within these subgroups did not allow for meaningful subgroup analyses. Consequently, no clear emerging pattern could be identified.

## CONCLUSION

In LMIC subnational cohorts, large-for-gestational-age newborns, particularly those above the 97th percentile, are associated with increased neonatal mortality. Macrosomia showed smaller and less robust associations across analyses, reflecting substantial uncertainty. Beyond expanding antenatal, intrapartum, and neonatal strategies to prevent and manage high-risk births, strengthened population-level surveillance, routine monitoring of birth weight trends, and timely collection of updated data are critical to identifying emerging patterns and tracking progress toward reducing avoidable neonatal deaths. Moreover, although LGA prevalence is currently low in these settings and the contribution of large newborns to neonatal mortality is much smaller than that of small newborns, integrating these measures into maternal and newborn health programs will be essential to accelerate progress toward global newborn survival goals.

## Author contribution

The Vulnerable Newborn Measurement Collaboration at Johns Hopkins University is led by E.A.H. F.K.-S. conceived the study and developed the analysis plan. All authors contributed to the study protocol and analysis approach. F.K.S., B.B.F., D.J.E., and E.A.H. carried out the statistical analyses. The draft of the manuscript was prepared by F.K.-S. with inputs of B.B.F., J.U., S.S., D.J.E., L.S.I, E.O.O., H.B., J.K., A.C.C.L. R.E.B., and E.A.H. All authors contributed to the revisions and approved the final manuscript.

## Acknowledgement

We thank all women and families included in datasets and all members of the study teams. We thank all relevant funders for their investments in enabling the input data for each of the included studies.

## Details of Ethics Approval

The Vulnerable Newborn Measurement Collaboration received ethical clearance from the Institutional Review Board of the John Hopkins Bloomberg School of Public Health (IRB No: 28757, approval date: 18 April 2024). Ethical approvals for the primary studies were obtained from all 23 countries’ teams from their respective ethical bodies.

## Data Availability Statement

The data that support the findings of this study are available on request from the corresponding author. The data are not publicly available due to privacy or ethical restrictions

