## Supplemental files for "Neonatal mortality risk of large-for-gestational age and macrosomic live births in low- and middle-income subnational birth cohorts: An individual participant meta-analysis (2000-2017)"

**ADDITIONAL FILE**

**Table S1: Characteristics of the studies**

| **Study ID** | **Period of data collection** | **Total live births** | **Total lost-to-follow-up** | **Neonatal deaths per 1000 livebirths (total sample)** | **Gestational age measurement**  **(% of sample for both LMP/ultrasound or timing of ultrasound)** | **Birthweights imputed** | **Setting** | **Study Design** | **Population represented** |
| --- | --- | --- | --- | --- | --- | --- | --- | --- | --- |
| **Eastern Asia** | | | | | | | | | |
| China 2002 (1) | 2002 – 2006 | 4,697 | 0 | 15.5 | LMP collected during pregnancy surveillance | Imputation of missing weights | Rural Shaanxi Province | Cluster RCT of antenatal supplementation with micronutrients interventions | Population based recruitment of all pregnant women in study area |
| **Latin America** | | | | | | | | | |
| Brazil 2015 (2) | 2015 | 4,275 | 0 | 8.7 | Combination of ultrasound (80%) and LMP (20%) | Imputation of missing weights | Urban Pelotas City | Prospective observational cohort study | Longitudinal birth cohort of all children delivered in the 5 maternity hospitals in the city of Pelotas |
| **sub-Saharan Africa** | | | | | | | | | |
| Burkina Faso 2004 and Burkina Faso 2006 (3,4) | 2004 – 2006  2006 – 2008 | 2,562 | 147 | 20.3 | Combination of ultrasound (74%) and LMP (26%) | Imputation of missing weights | Rural Hounde | RCT of multiple micronutrient supplementation  RCT of maternal fortified food supplementation | Prospective, community-based cohort |
| Malawi 2003 and Malawi 2011 (5,6) | 2003 – 2006  2011 – 2012 | 2,540 | 33 | 28.3 | Ultrasound in 1^st^ trimester (100%) | Recalibration of all weights to time at delivery & imputation of missing | Rural Mangochi District | RCT of sulfadoxine-pyrimethamine and azithromycin in pregnancy  RCT of lipid-based nutrient supplement | Facility-based ANC clinic recruitment of pregnancies |
| Tanzania 2008 (7) | 2008 – 2010 | 915 | 0 | 28.1 | Ultrasound (100%) | Exclude weights measured >72 hours | Rural Korogwe | Prospective observational cohort studies | - Population-based recruitment of women preconception or in the 1st trimester of pregnancy - Facility-based ANC clinic recruitment of pregnancies |
| Tanzania 2014 (8) | 2014 – 2016 | 426 | 0 | 29.5 | Ultrasound (100%) | Exclude weights measured >72 hours | Rural Korogwe | Prospective observational cohort studies |  |
| Zambia 2011 (9) | 2011 – 2013 | 37,856 | 1156 | 14.4 | LMP collected during pregnancy surveillance | Recalibration of all weights to time at delivery & imputation of missing | Urban and rural areas of Southern Province | RCT of chlorhexidine application for umbilical cord disinfection | Facility-based ANC clinic recruitment of pregnancies |
| Zambia 2015 (10) | 2015 – 2017 | 11,016 | 0 | 12.3 | LMP collected during pregnancy surveillance | Imputation of missing weights | Urban Lusaka | Prospective observational cohort study | Population based recruitment of all pregnant women in study area |
| **Southern Asia** | | | | | | | | | |
| Bangladesh 2011 (11,12) | 2011 – 2014 | 21,227 | 874 | 45.1 | LMP collected during pregnancy surveillance | Recalibration of all weights to time at delivery & imputation of missing | Rural Sylhet | Prospective observational cohort study | Population-based recruitment of all pregnant women in study area |
| Bangladesh 2012 (13) | 2012 – 2015 | 7698 | 0 | 15.5 | LMP collected during pregnancy surveillance | Recalibration of all weights to time at delivery & imputation of missing | Rural Sylhet | RCT of antenatal screening and treatment of genitourinary tract infections | Population-based recruitment of all pregnant women in study area |
| Bangladesh 2014 | 2015 – 2017 | 2858 | 0 | 18.9 | Ultrasound (100%) | Recalibration of all weights to time at delivery & imputation of missing | Rural Sylhet | Cohort study to assess late pregnancy biometry for gestational age | Population-based recruitment of all pregnant women in study area |
| India 2000 (14) | 2000 – 2001 | 5,890 | 7 | 33.3 | LMP collected during pregnancy surveillance | Recalibration of all weights to time at delivery & imputation of missing | Rural Tamil Nadu | RCT of newborn Vitamin A supplementation | Population-based recruitment of all pregnant women in study area |
| Nepal 2002 (15,16) | 2002 – 2005 | 23,665 | 88 | 30.0 | LMP collected during pregnancy surveillance | Recalibration of all weights to time at delivery & imputation of missing | Rural Sarlahi District | Cluster RCT of newborn skin-umbilical cord cleansing with chlorhexidine | Population-based recruitment of all pregnant women in study area |
| Nepal 2010 (17) | 2010 – 2017 | 31,116 | 1,157 | 31.9 | LMP collected during pregnancy surveillance | Recalibration of all weights to time at delivery & imputation of missing | Rural Sarlahi District | Cluster RCT of newborn massage with sunflower seed oil | Population-based recruitment of all pregnant women in study area |

LMP: Last menstrual period; RCT: randomized control trial

### Table S2: Sensitivity analyses of the association between macrosomia, large-for-gestational-age, and neonatal mortality across gestational periods using generalized linear mixed models

| Analysis Subset | Odds Ratio (OR) | 95% CI Lower | 95% CI Upper | P-Value |
| --- | --- | --- | --- | --- |
| LGA 90 (Overall) | 2.57 | 2.28 | 2.88 | <0.001 |
| LGA 90 (early mortality) | 2.89 | 2.54 | 3.28 | <0.001 |
| LGA 90 (late mortality) | 1.43 | 1.08 | 1.92 | 0.01 |
| LGA 90 (Term) | 0.86 | 0.56 | 1.30 | 0.46 |
| LGA 97 (Overall) | 3.34 | 2.92 | 3.82 | <0.001 |
| LGA 97 (early mortality) | 3.88 | 3.36 | 4.49 | <0.001 |
| LGA 97 late mortality | 1.62 | 1.14 | 2.30 | 0.007 |
| LGA 97 (Term) | 1.23 | 0.65 | 2.32 | 0.52 |
| Macrosomia (Overall) | 0.80 | 0.49 | 1.32 | 0.39 |
| Macrosomia (early mortality) | 0.72 | 0.39 | 1.31 | 0.20 |
| Macrosomia (late mortality) | 1.15 | 0.47 | 2.80 | 0.76 |
| Macrosomia ≥4500g (Overall) | 1.14 | 0.36 | 3.56 | 0.82 |

### OR: Odds Ratio, CI: confidence interval. LGA: large-for-gestational-age (90 percentile, 97 percentile), Macrosomia is defined as birth weight ≥4000 g. All analyses were based on imputed data.

### Table S3: Sensitivity analyses of the association between macrosomia, large-for-gestational-age, and neonatal mortality across gestational periods using a beta–binomial model

| Analysis Subset | RR | 95% CI Lower | 95% CI Upper | P-Value |
| --- | --- | --- | --- | --- |
| LGA > 90 (Overall) | 1.88 | 1.18 | 2.99 | 0.008 |
| LGA > 90 (early morality) | 2.04 | 1.22 | 3.41 | 0.007 |
| LGA > 90 (late mortality) | 1.29 | 0.87 | 1.92 | 0.20 |
| LGA > 97 | 2.48 | 1.50 | 4.12 | <0.001 |
| LGA > 97 (early mortality) | 2.74 | 1.57 | 4.77 | <0.001 |
| LGA > 97 (late mortality) | 1.41 | 0.90 | 2.21 | 0.13 |
| Macrosomia (Overall) | 0.86 | 0.45 | 1.65 | 0.65 |
| Macrosomia (early mortality) | 0.83 | 0.40 | 1.71 | 0.61 |
| Macrosomia (late mortality) | 0.97 | 0.38 | 2.49 | 0.96 |
| Macrosomia ≥4500g (Overall) | 1.15 | 0.36 | 3.66 | 0.81 |

### RR: risk ratio, CI: confidence interval, LGA: large-for-gestational-age (90th percentile, 97th percentile). Macrosomia is defined as birth weight ≥4000 g. All analyses were based on imputed data.

| **A** | 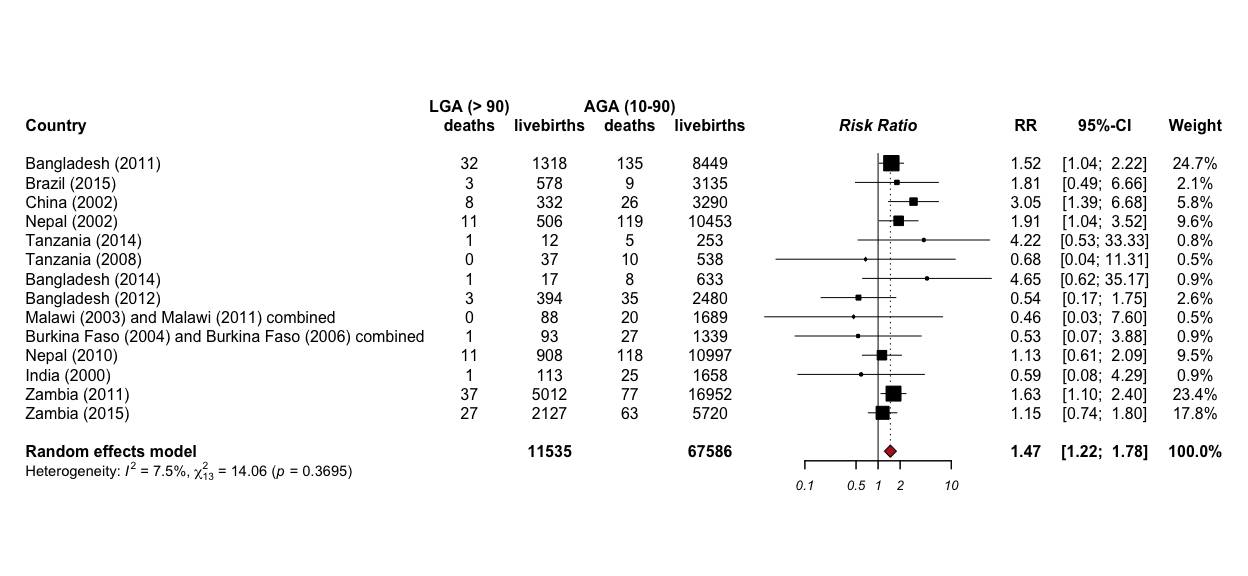 |
| --- | --- |

**Figure S1.** Neonatal mortality risk (RR) in the LGA >90th percentile as calculated using non-imputed data with Haldane correction. AGA, appropriate for gestational age; LGA, large for gestational age; RR, relative risk.

|  | 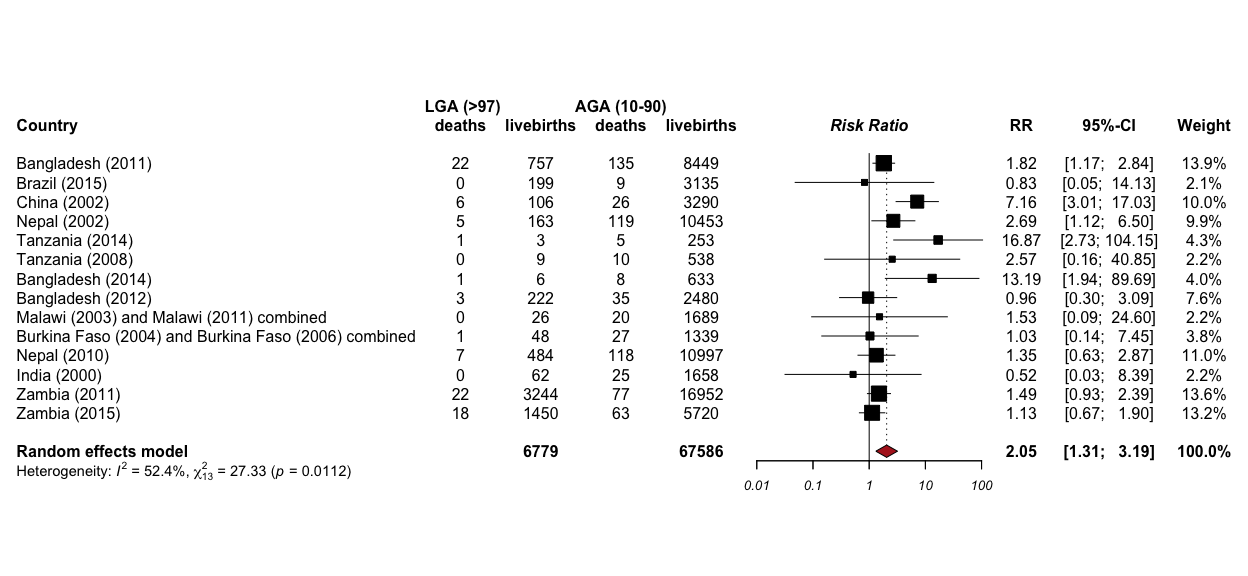 |
| --- | --- |

**Figure S2.** Neonatal mortality risk in the LGA >97th percentile as calculated using non-imputed data with Haldane correction . AGA, appropriate for gestational age; LGA, large for gestational age; RR, relative risk.

| **A** | 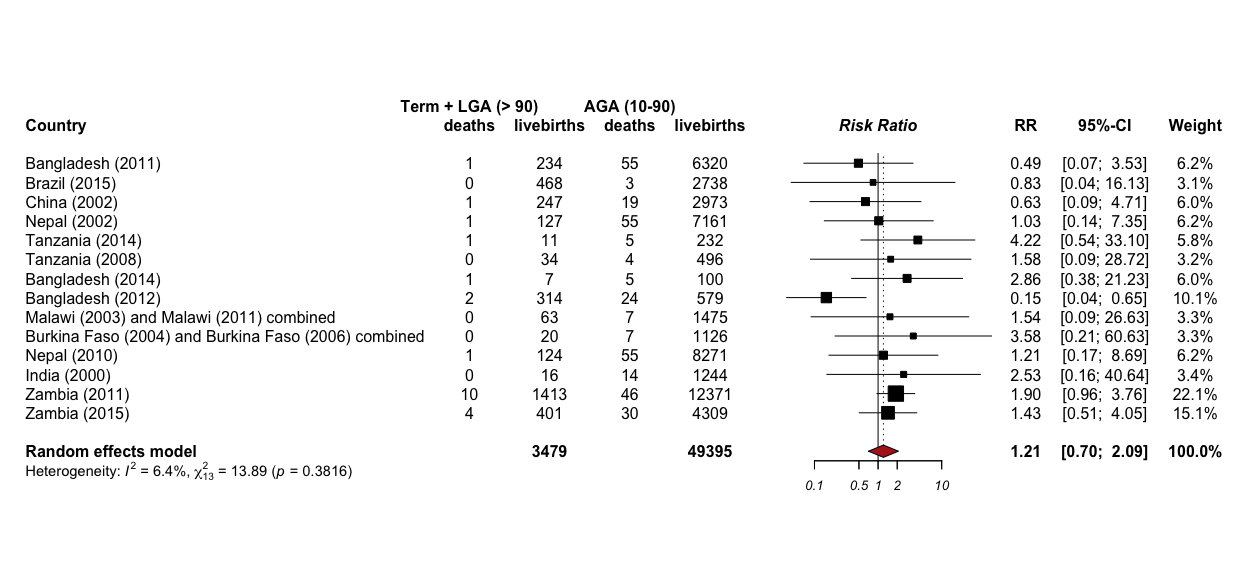 |
| --- | --- |

**Figure S3.** Neonatal mortality risk in LGA >90th percentile term neonates, calculated from non-imputed data with the Haldane correction. AGA, appropriate for gestational age; LGA, large for gestational age; RR, relative risk.

|  | 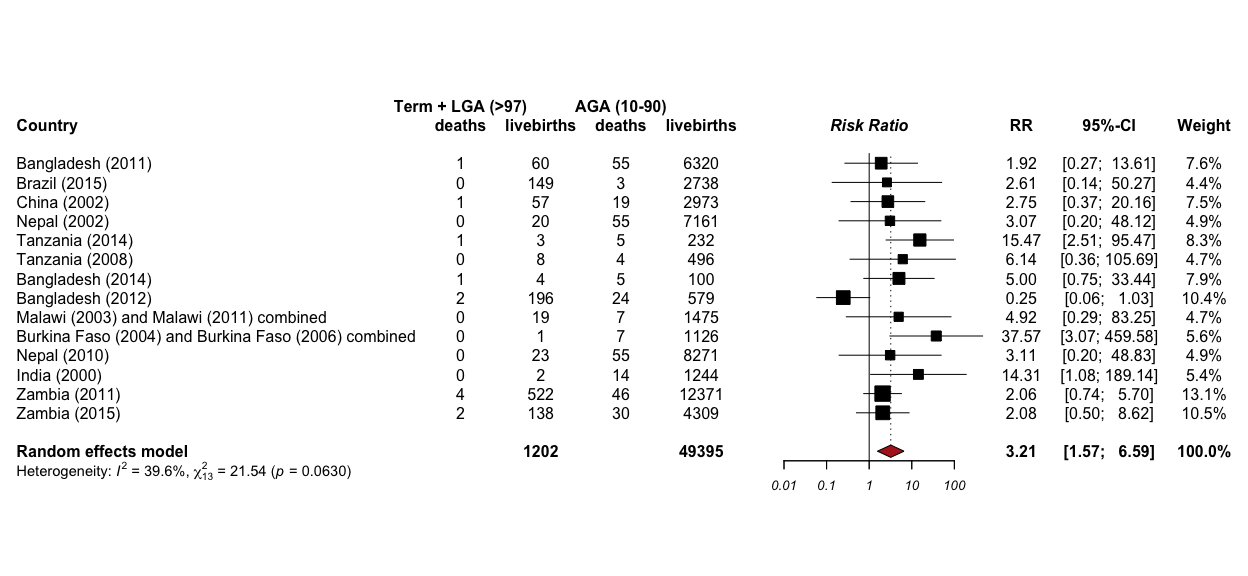 |
| --- | --- |

**Figure S4.** Neonatal mortality risk in LGA >97th percentile term neonates, calculated from non-imputed data with the Haldane correction. AGA, appropriate for gestational age; LGA, large for gestational age; RR, relative risk.

| **A** | 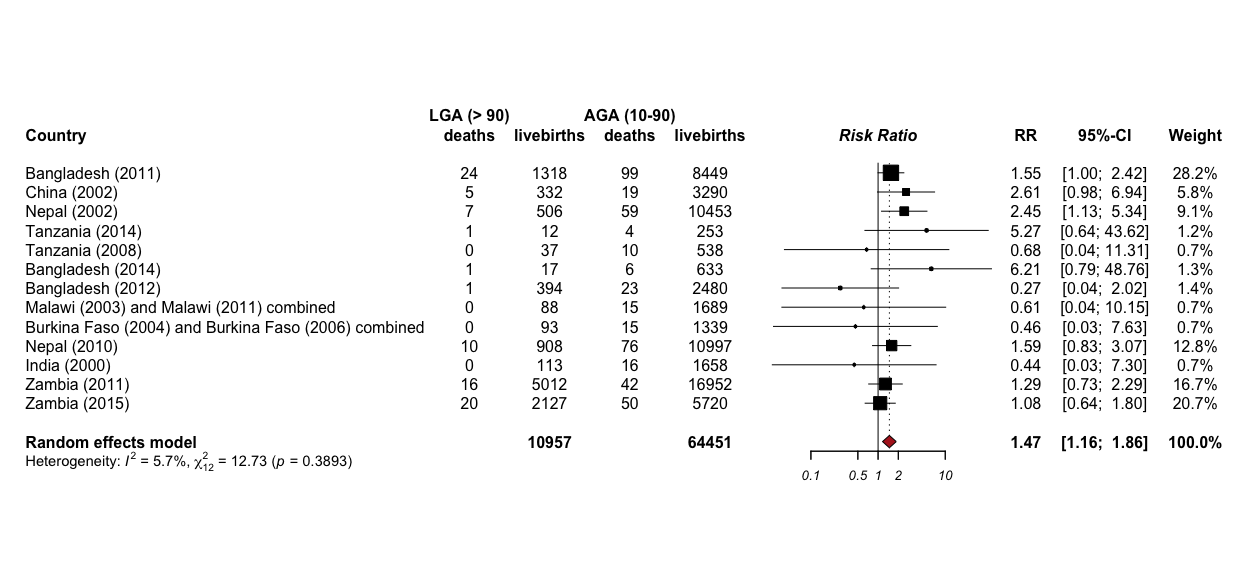 |
| --- | --- |

**Figure S5.** Early neonatal mortality risk in the LGA >90th percentile as calculated from **non-imputed** data with the Haldane correction. AGA, appropriate for gestational age; LGA, large for gestational age; RR, relative risk.

|  | 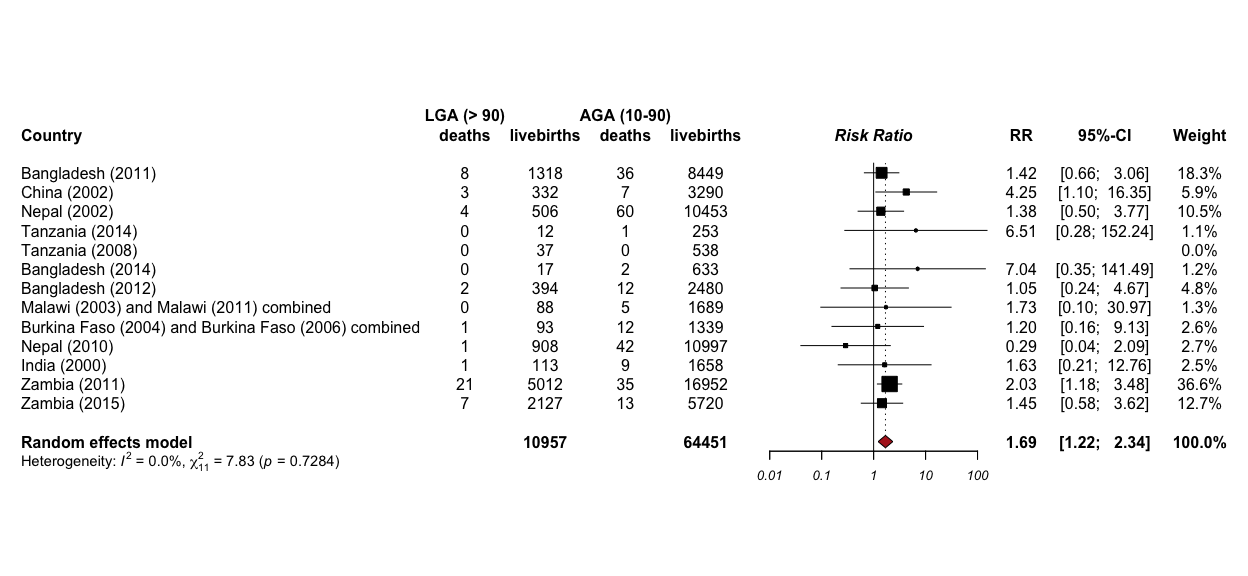 |
| --- | --- |

**Figure S6.** Late neonatal mortality risk in the LGA >90th percentile as calculated from **non-imputed** data with the Haldane correction. AGA, appropriate for gestational age; LGA, large for gestational age; RR, relative risk.

|  | 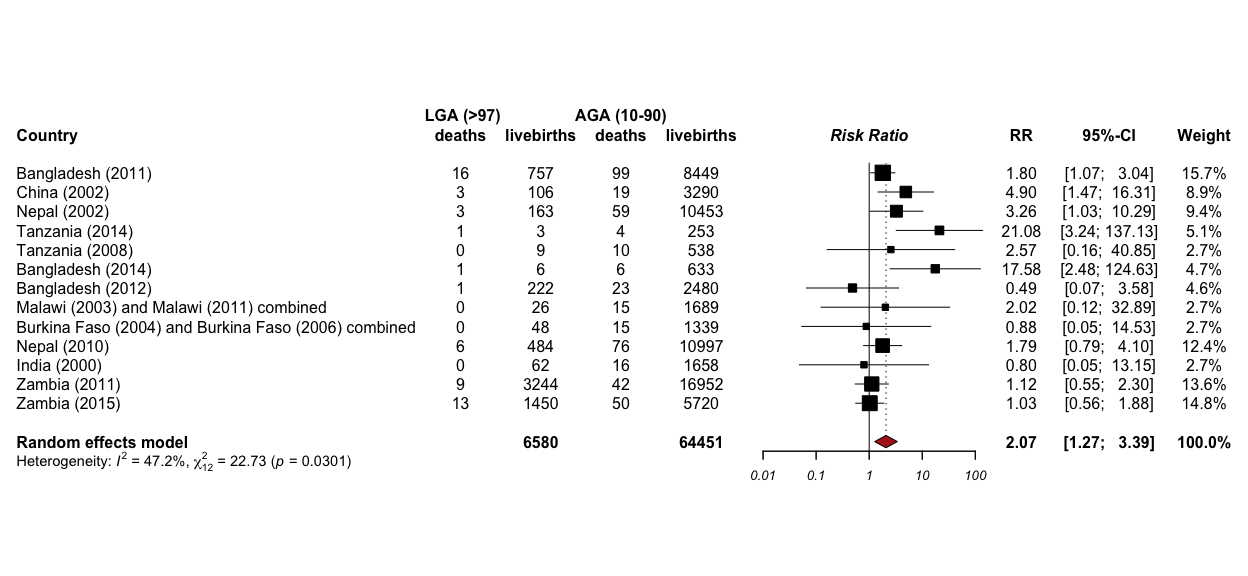 |
| --- | --- |

**Figure S7.** Early neonatal mortality risk in the LGA >97th percentile as calculated from **non-imputed** data with the Haldane correction. AGA, appropriate for gestational age; LGA, large for gestational age; RR, relative risk.

|  | 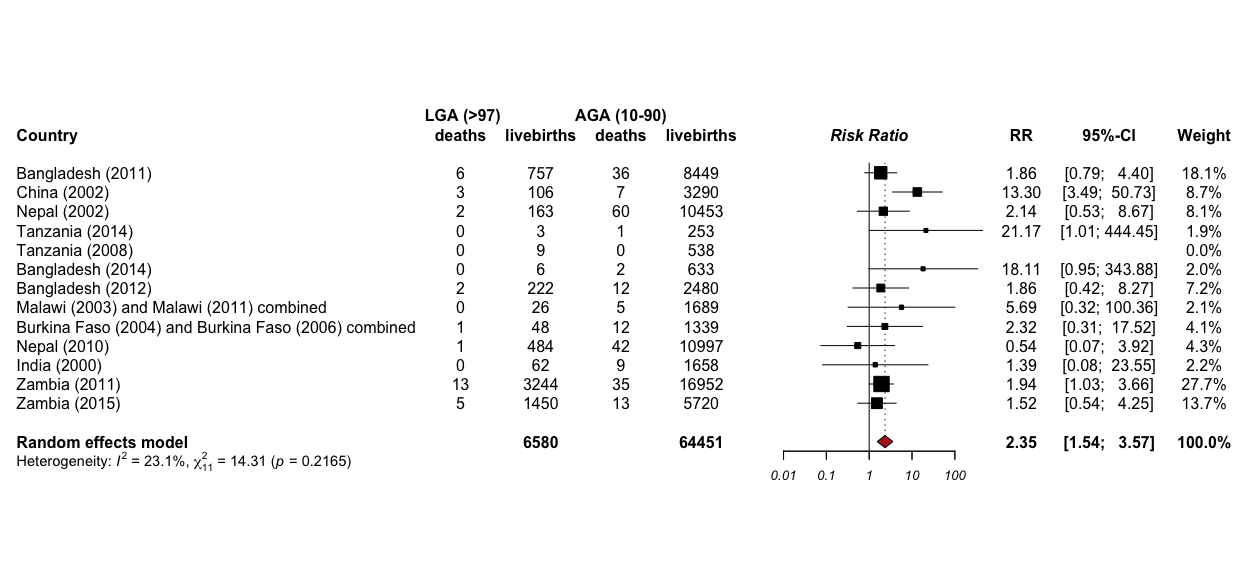 |
| --- | --- |

**Figure S8.** Late neonatal mortality risk in the LGA >97th percentile as from **non-imputed** data with the Haldane correction. AGA, appropriate for gestational age; LGA, large for gestational age; RR, relative risk.

|  | 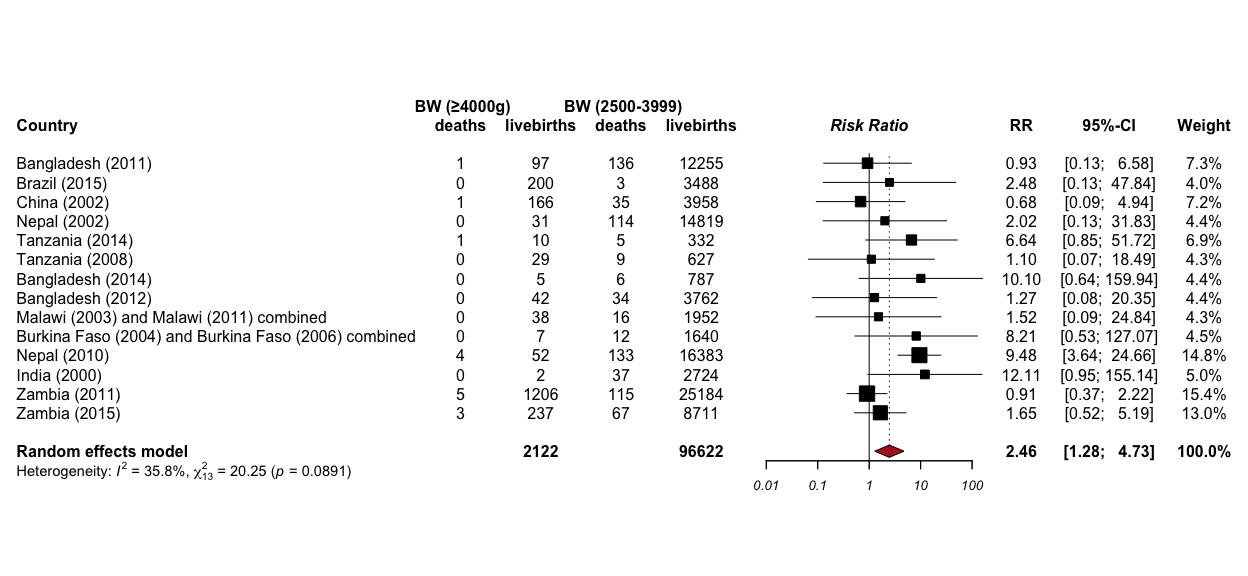 |
| --- | --- |

**Figure S9.** Neonatal mortality risk in infants with macrosomia >4000 g as calculated from **non-imputed** data with the Haldane correction. BW: birth weight. RR: risk ratio

|  | 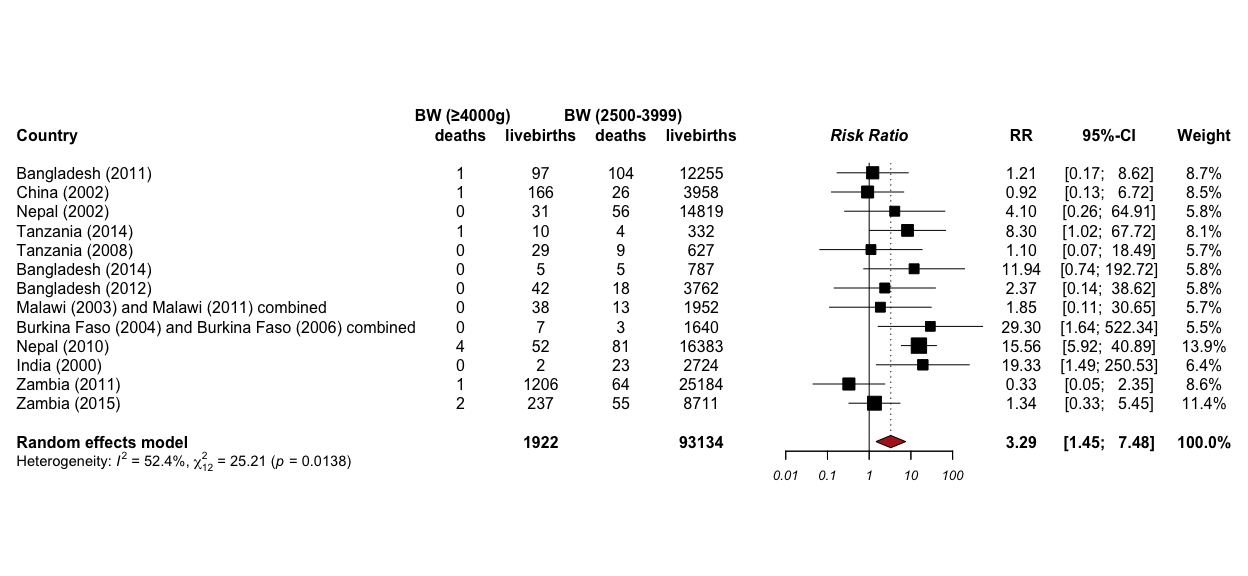 |
| --- | --- |

**Figure S10.** Early neonatal mortality risk in infants with macrosomia >4000 g as calculated from **non-imputed** data with the Haldane correction. BW, birth weight. RR, risk ratio

| **A** | 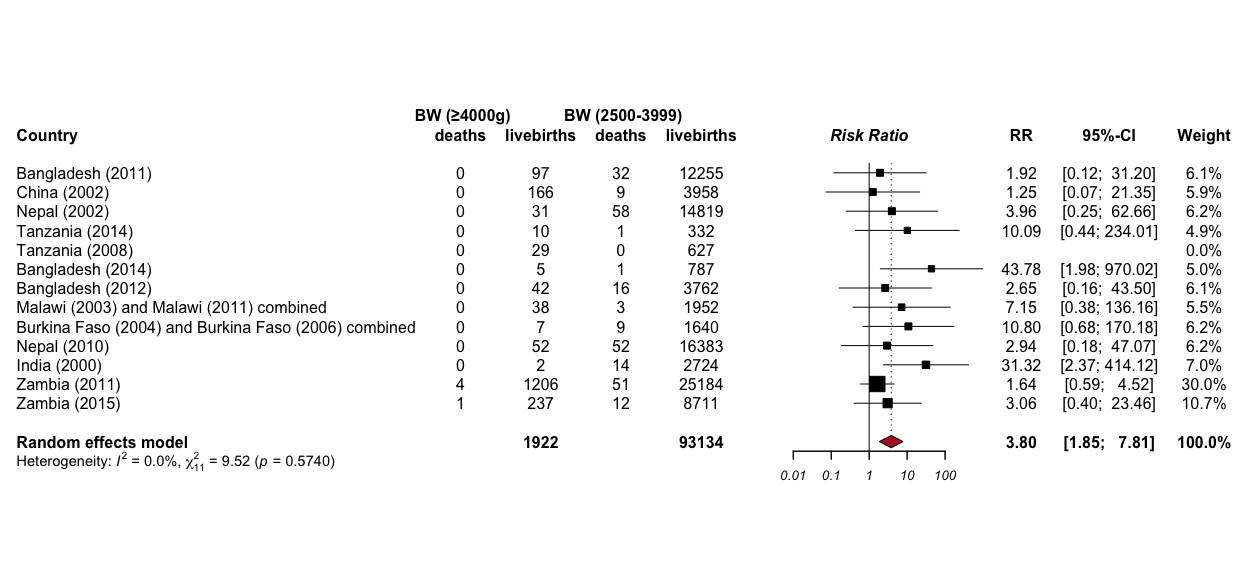 |
| --- | --- |

**Figure S11.** Late neonatal mortality risk in infants with macrosomia >4000 g as calculated from **non-imputed** data with the Haldane correction. BW, birth weight. RR, risk ratio
